# Benchmarking the AI-based diagnostic potential of plasma proteomics for neurodegenerative disease in 17,187 people

**DOI:** 10.1101/2025.06.27.25330344

**Authors:** Lijun An, Alexa Pichet-Binette, Ines Hristovska, Gabriele Vilkaite, Xiao Yu, Romina Zendehdel, Zijian Dong, Bart Smets, Rowan Saloner, Shinya Tasaki, Ying Xu, Varsha Krish, Farhad Imam, Shorena Janelidze, Danielle van Westen, the Global Neurodegenerative Proteomics Consortium (GNPC), Erik Stomrud, Christopher D. Whelan, Sebastian Palmqvist, Rik Ossenkoppele, Niklas Mattsson-Carlgren, Oskar Hansson, Jacob W. Vogel

## Abstract

Co-pathology is a common feature of neurodegenerative diseases that complicates diagnosis, treatment and clinical management. However, sensitive, specific and scalable biomarkers for in vivo pathological diagnosis are not available for most neurodegenerative neuropathologies. Here, we present ProtAIDe-Dx, a deep joint-learning model trained on 17,187 patients and controls (Age=70.3±11.5, 53.2% of Female) that uses plasma proteomics to provide simultaneous probabilistic diagnosis across six conditions associated with dementia in aging. ProtAIDe-Dx achieves cross-validated balanced classification accuracy of 70%-95% and AUCs > 78% across all conditions. The model’s diagnostic probabilities highlighted subgroups of patients with co-pathologies, and were associated with pathology-specific biomarkers in an external memory clinic sample, even among cognitively unimpaired people. Model interpretation revealed a suite of protein networks marking shared and specific biological processes across diseases, and identified novel and previously described proteins discriminating each diagnosis. ProtAIDe-Dx significantly improved biomarker-based differential diagnosis in a memory clinic sample, pinpointing proteins leading to diagnostic decisions at an individual level. Together, this work highlights the promise of plasma proteomics to improve patient-level diagnostic work-up with a single blood draw.

## Introduction

The last five years have seen multiple breakthroughs in the treatment of neurodegenerative diseases. Early disease modifying therapies have emerged for Alzheimer’s disease (AD) ^1,2^, and highly promising drug candidates are currently in clinical trials for AD ^3^, Parkinson’s disease (PD) ^4^ and amyotrophic lateral sclerosis (ALS) ^5^. However, differential diagnosis and disease comorbidity continue to pose considerable challenges in these treatment efforts. Misdiagnosis rates are around 25-30% even in specialized dementia clinics, and can exceed 50% in primary care ^6–8^. Meanwhile, comorbidity is common in aging, with 70% of patients 80 years or older harboring multiple neurodegenerative pathologies simultaneously ^9^. Misdiagnosis can make it difficult to select the right patients for a drug trial ^10^, while comorbid neuropathologies can mask the positive effects of a putative therapy ^11,12^. Once such treatments do become available, misdiagnosis and comorbidity can both lead to treatment mismanagement ^13^. With the rapid pace at which promising new drugs are being tested, there is an urgent need for powerful tools for diagnosis and precise identification of underlying comorbid pathologies.

The first step toward mitigating the issues presented by misdiagnosis and comorbidity is the development of biomarkers to identify underlying neurodegenerative pathology with high specificity. As an example, highly sensitive and specific fluid ^14–18^ and imaging ^19,20^ biomarkers for AD have emerged and have become instrumental in the recent development of disease modifying therapies. AD diagnostic capabilities have expanded even further as sensitive tests for underlying AD pathology are now possible using only blood plasma ^21,22^. Blood-based biomarkers have the potential to be highly accessible, inexpensive and minimally invasive, and the emergence of blood-based biomarkers for AD could facilitate accurate AD diagnosis even in primary care in the near future ^7^. Despite the success of AD biomarkers, scalable sensitive and specific biomarkers for other neurodegenerative diseases are lacking, and diagnosis can only be made with high confidence at autopsy.

Given the diversity of underlying diseases that patients commonly present with at memory clinics ^23,24^, and given the substantial effects of comorbid pathology on prognosis ^25,26^, a multi-disease diagnostic test would be of great benefit to researchers, clinical neurologists and their patients. Plasma proteomics represent a promising tool toward this aim, allowing robust surveillance of thousands of potential biomarkers and relevant functional effectors with a single blood draw ^27–29^. While still a new and rapidly evolving tool, plasma proteomics has already shown promise in the simultaneous prediction of multiple health outcomes ^30–32^ and in the monitoring of the integrity of multiple organs and critical biological systems ^33^. Beyond their capacity as potential biomarkers, circulating proteins further provide a view into complex molecular dynamics as they play out in living humans, providing novel biological insight into disease processes ^34–37^.

Despite great promise, plasma proteomics data is not without its challenges. Proteomics data is high rank ^38^, comes with burdensome technological artifacts ^39,40^, and likely represents complex nonlinear interactions ^41^. In addition, the blood-brain barrier limits the number of brain-expressed proteins relevant in neurological conditions that can be detected in blood ^42,43^(although molecular processes governing selective protein and macromolecule transport across the BBB remain incompletely understood ^44,45^). Sophisticated modeling techniques such as artificial intelligence (AI) will likely be needed to synthesize clinically useful disease signatures from this rich and complex data^46^. Although some studies have leveraged population-based cohorts ^47–50^, the potential of AI-proteomics has not yet been benchmarked in large neurodegenerative cohorts. In the present study, we attempt to overcome these limitations by applying AI to the Global Neurodegenerative Proteomics Consortium (GNPC) v1.3MS dataset, the largest neurodegenerative disease plasma proteomics dataset to date. We present a model called Proteomics-based AI for Dementia Diagnosis (ProtAIDe-Dx), a deep multi-task architecture to resolve differential and multiple neurodegenerative diagnoses with a single blood draw (Fig. 1A). ProtAIDe-Dx is based on a joint-learning framework that simultaneously resolves multiple diagnostic tasks, thus learning shared signatures across diseases from a reduced set of disease-relevant proteins. ProtAIDe-Dx provides probabilistic diagnostic information across five of the most common incapacitating neurological conditions in old age – AD, PD, stroke, ALS and frontotemporal dementia (FTD). Diagnostic predictions are based on nonlinear proteomic embeddings that can be extracted and used for few-shot transfer learning to related tasks and unseen datasets. We report the performance of ProtAIDe-Dx on multi-diagnosis prediction, evaluate its potential in identifying co-pathology, test its capability in differential diagnosis compared to other clinical markers, and explore putative molecular networks contributing to its predictions. Finally, we present a proof-of-concept using ProtAIDe-Dx for personalized and interpretable diagnostic testing in a clinical scenario. With this approach, we hope to set a benchmark for future plasma based multi-disease neurodegenerative diagnostic tools.

## Results

A subsample of 17,187 participants with SomaLogic 7k proteomics, sampled across 19 contributing sites, was selected from the GNPC v1.3MS dataset for subsequent analysis (see Methods). Table S1 describes sample and demographics across each contributing site, while Table S2 lists the frequency of six conditions across each site, namely AD, PD, FTD, ALS, previous stroke/transient ischemic attack (TIA), and cognitively unimpaired. Given the high prevalence of vascular dementia (second only to AD^51^) but the lack of vascular dementia diagnoses in GNPC, the stroke/TIA group was chosen as a representation of patients with documented cerebrovascular disease. The total sample summed to 6,693 recruited controls and 5,772 patients. Of the patients, 2,249 were diagnosed with AD, 2,287 were diagnosed with PD, 435 with ALS, 175 with FTD and 317 with stroke/TIA. 309 patients had two or more diagnoses. 4,722 participants were neither labeled as Controls nor diagnosed with the five diseases above: 3,116 were MCI-SCI, 1,064 had dementia (based on MMSE or CDR), and 542 had unknown status. None of these 4,722 cases were included in model development, but served as independent test groups (see Methods). We applied the ProtAIDe-Dx model to this sample, an architecture capable of generating several features of interest simultaneously: binary diagnosis of each condition, probabilities of each diagnosis, and joint embeddings representing low-dimension, non-linear protein combinations used by the model for diagnosis. Fig. 1A describes our overall workflow and how each of these model features is used. The model architecture of ProtAIDe-Dx was set as a multiple layer perceptron (MLP), where optimal layers and nodes per layer were chosen by hyperparameter searching on nested validation folds (see Methods). We specifically chose a multi-task, joint learning approach (as opposed to a multi-class classification task) to allow the model to signal disease co-pathology (i.e. positive for multiple disorders and probabilities for each disorder).

**Figure 1.**
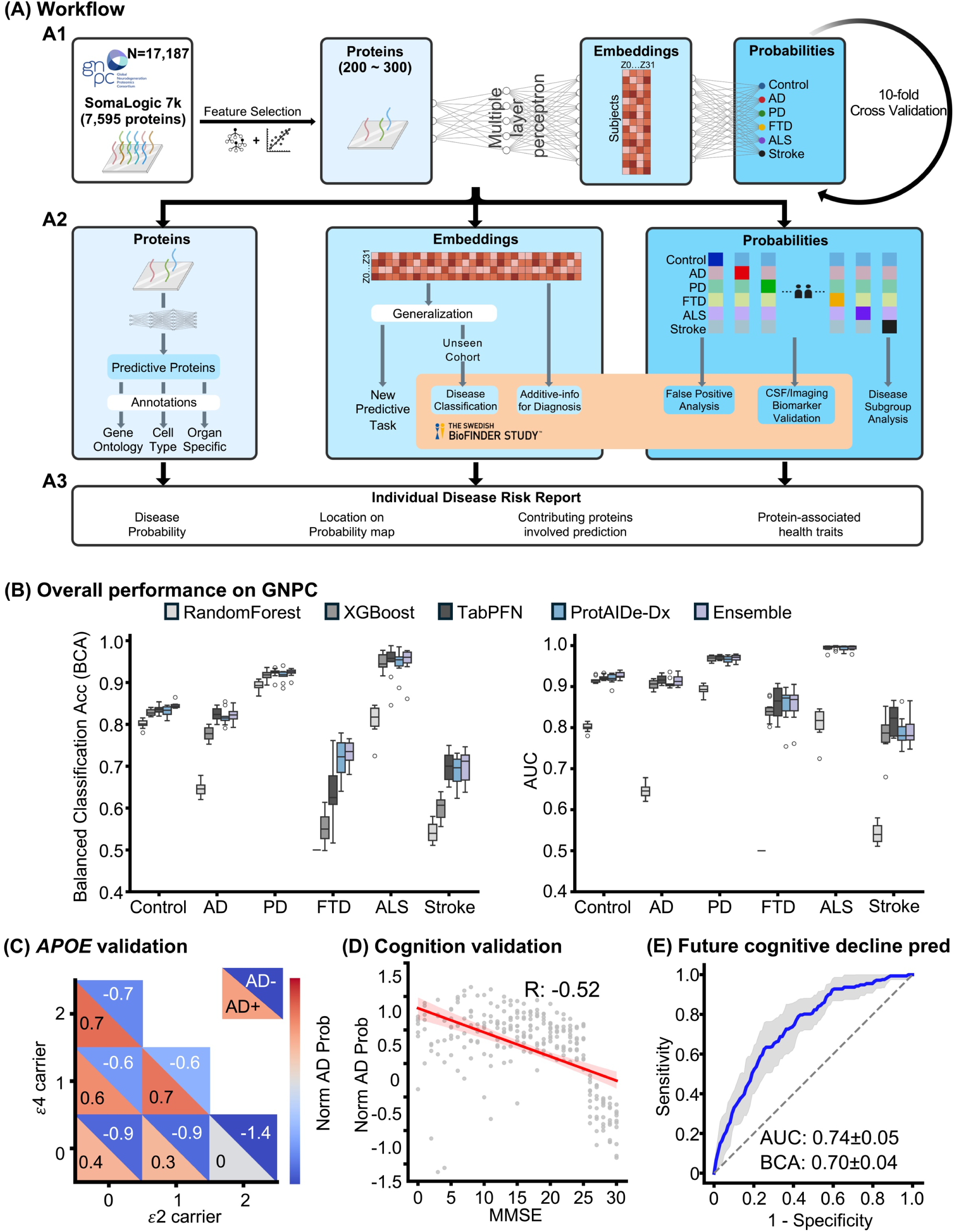
Workflow and overall performances of ProtAIDe-Dx on GNPC. (A) Project workflow. (A1) Model development on GNPC with 10-fold cross-validation procedure. The joint learning framework allows learning of several neurodegenerative classification tasks altogether, facilitating shared information during training. (A2) Model evaluation on GNPC (blue panels) and BioFINDER-2 cohort (orange panel). Model outputs include not just classifications, but also probabilities for each class, and contributing proteins and embeddings can be probed to better understand model choices. (A3) Individual neurological disease risk report based on developed model. (B) Overall model performances on GNPC. Left: Balanced classification accuracy (BCA) scores for each diagnostic task; Right: AUC score for each diagnostic task. (C) Normalized AD probabilities stratified by different *APOE* ε2/ε4 groups, shown in participants with (lower diagonal) and without (upper diagonal) AD diagnosis. (D) Correlation between normalized AD probabilities and MMSE. (E) ROC curve of model generalization to a new task for predicting longitudinal clinical progression (from no cognitive impairment to future cognitive impairment).

### Joint learning improves multi-diagnostic prediction of neurodegenerative diagnosis from blood in unbalanced samples

We applied ProtAIDe-Dx to the GNPC sample, using 10-fold cross-validation stratified for each contributing site. Importantly, we used only proteomic information in the model – no site, demographic, cognitive or diagnostic information was used. We compared the diagnostic performance of ProtAIDe-Dx against multiple machine learning and state-of-the-art deep learning baselines, including Random Forest, XGBoost^52^, and TabPFN^53^. Random Forest and XGBoost are classical yet high-performing machine learning models on tabular data, which we trained independently for each task. TabPFN, in contrast, is a state-of-the-art transformer-based model pretrained on millions of synthetic datasets and optimized for binary classification; accordingly, we trained separate TabPFN models for each diagnostic task. ProtAIDe-Dx emerged as the best-performing model overall, while XGBoost was the best non–deep learning models (Fig. 1B). Due their highly disparate approaches, we anticipated that ProtAIDe-Dx and XGBoost might employ different classification strategies, and therefore might provide complementary information in an ensemble framework. To test this hypothesis, we also tested an ensemble model including aspects of both models.

ProtAIDe-Dx achieved a median balanced accuracy performance above 90% for ALS (95%) and PD (92%) classification, 83% for Control, 81% for AD, 72% for FTD, and 70% for stroke/TIA (Fig. 1B). ProtAIDe-Dx significantly outperformed Random Forest across all tasks, XGBoost in AD (FDR-corrected p = 5e-4), FTD (FDR-corrected p = 3e-4), and stroke (FDR-corrected p = 0.004) classification, and significantly outperformed TabPFN in FTD classification (FDR-corrected p = 0.047). With the exception of Random Forest, all models achieved AUCs >0.8 for all tasks other than stroke/TIA prediction and demonstrated comparable AUCs (Fig. 1B, Table S3, SuppData 1), though AUC alone might convey overoptimistic implications in some imbalanced classification scenarios^54,55^ (Fig. S1). We found that the ensemble model produced balanced accuracy scores and AUC that significantly outperformed ProtAIDe-Dx for Control and PD diagnosis. The ensemble model also significantly improved balanced accuracy scores over XGBoost for all tasks except ALS and PD prediction (Fig. 1B, Table S3, SuppData 1). In all, AI-based proteomic models showed impressive multi-disease diagnostic performance, and the deep, joint-learning framework provided by ProtAIDe-Dx afforded performance advantages over a canonical tree-based, single-task baseline, as well as comparable performance to state-of-the-art models.

As a sanity check, we extracted the probability of AD diagnosis across all individuals and compared them to factors known to be altered in AD, namely cognitive performance and *APOE* ε2 and ε4 allele carriage. In both patients diagnosed with AD and those without AD diagnosis, presence of more copies of the *APOE* ε4 carriage was associated with higher AD probabilities, and of more copies of the ε2 was associated with lower AD probabilities (Fig. 1C). Additionally, a strong negative correlation was observed between AD probabilities and MMSE (mini-mental status examination) score, a measurement of global cognition, indicating worse cognition associated with higher AD probabilities (Fig. 1D). These analyses suggest model-derived diagnostic probabilities can serve as continuous proteomic scores associated with indicators of disease progression. We further test this hypothesis in an external dataset with complete biomarkers below.

### Diagnostic prediction model generalizes to new disease-relevant tasks

Low-dimensional nonlinear proteomic embeddings were extracted from the last layer of the ProtAIDe-Dx model (Fig. 1A). Given ProtAIDe-Dx’s joint-learning architecture, these embeddings should represent a compressed representation of plasma proteomic data optimized toward tasks related to neurological diseases. To test this hypothesis, we used these embeddings to generalize the ProtAIDe-Dx model to a task that ProtAIDe-Dx was not trained specifically for, namely prediction of longitudinal clinical progression in healthy controls (see Methods). The performance of this model in predicting individuals who progressed from unimpaired to impaired (i.e. from clinical dementia rating [CDR] 0 to CDR 0.5 or 1; N=218), compared to those that remained stably at CDR 0 over time (N=1,445), reached a balanced classification accuracy of 70% and an AUC of 74% (Fig. 1E). Notably, this model generalization was achieved simply by transferring the embeddings with a simple logistic regression model, a process that requires minimal CPU power and does not require any GPUs given – important that global GPU inequality is expected to persist at least in the near future. These results support ProtAIDe-Dx as a flexible and extensible model for neurodegenerative disease-related tasks.

**Figure 2.**
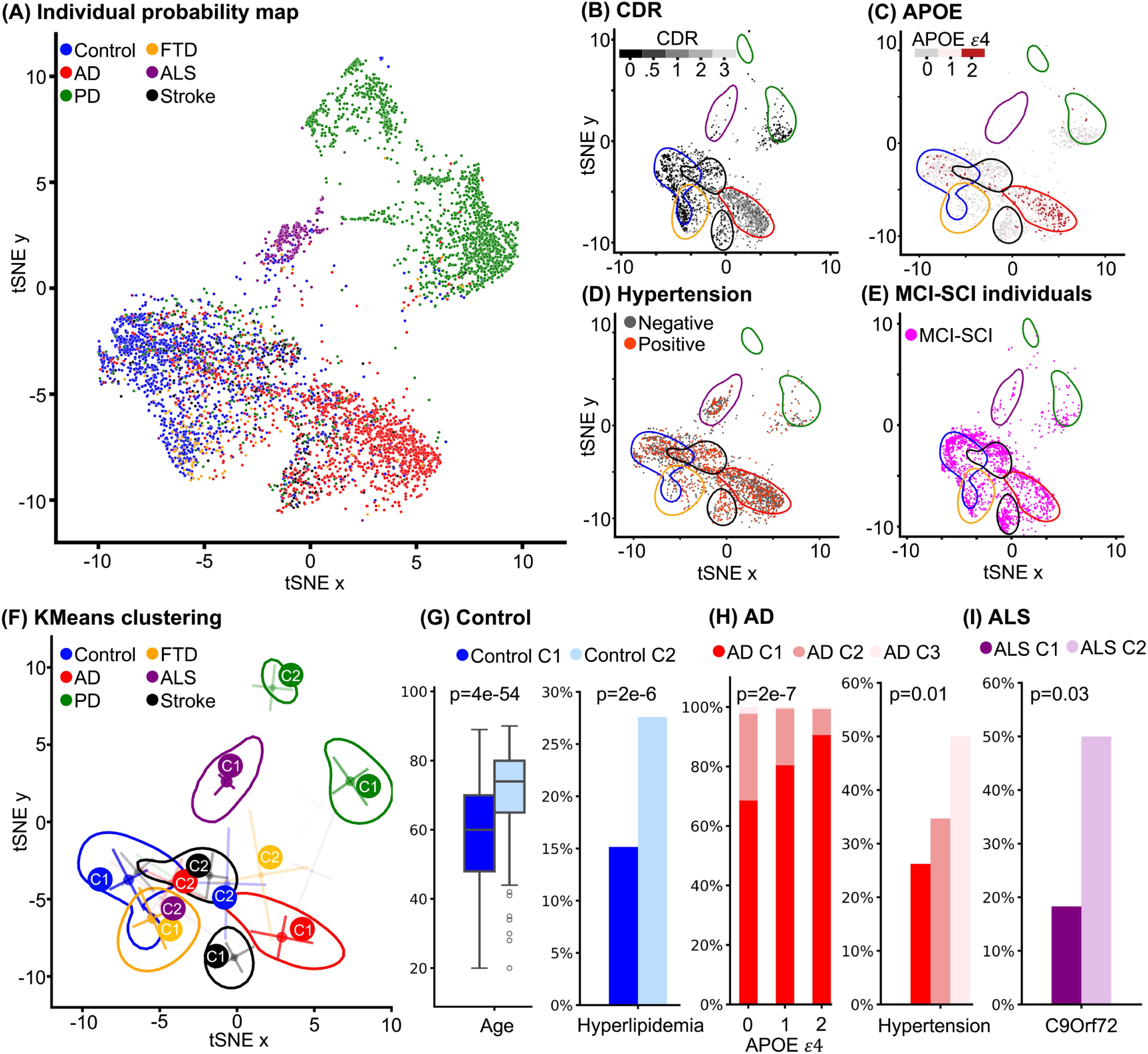
Diagnostic probability map derived by ProtAIDe-Dx reveal disease heterogeneity. (A) Participants from the test set were selected for tSNE to reduce dimensionality from six (predicted class probabilities) to two (nonlinear disease probability axes), colored by clinical diagnosis. (B) - (E) Participants were projected onto the fitted 2-dimensional diagnostic probability map, colored by variable values. (F) KMeans clustering within each diagnostic category based on their disease probabilities, where the top two clusters were annotated. The contour was drawn based on Gaussian kernel density estimation with a threshold of 0.01. (G) - (I) Distribution of variables varied across the Control, AD, and ALS clusters (see Fig. S4 for further detail on disease clusters).

### Diagnostic probabilities reveal disease heterogeneity and co-pathology

ProtAIDe-Dx provides probabilities of each condition for each individual. To visualize the overall landscape of how individuals are distributed based on probabilities, we projected all individuals into a two-dimensional nonlinear embedding based on their vector of disease probabilities (Fig. 2A). As expected, individuals naturally clustered based on their clinical diagnosis. PD and ALS cases formed their own separate islands (reflecting the ease with which these conditions were classified). In contrast, a central island emerged with AD and cognitive normal individuals on either end of the spectrum, and offshoots from the center showing higher density of FTD cases on one side, whereas stroke/TIA cases appeared more sparsely, located both near the island’s center and along one side of the spectrum. Importantly, these islands were driven primarily by etiology, and not by contributing site, with the exception of one of the two PD islands which consisted entirely of PD patients from Site U (Fig. S2). Common phenotypic data in the GNPC distributed in expected patterns across the embeddings, with worse cognitive impairment and more *APOE* ε4 carriers in AD regions, less ε4 carriers in PD and ALS regions, and more hypertension in the stroke/TIA region (Fig. 2B, C, D).

Notably, patients diagnosed with subjective cognitive decline (SCD) or mild cognitive impairment (MCI) were not used to train ProtAIDe-Dx, due to the etiology being less clear in these early phases of impairment. We therefore used ProtAIDe-Dx to predict etiological diagnoses for these SCD and MCI cases (N=3,116) and projected these cases onto the diagnostic embedding (Fig. 2E). Interestingly, the cases were distributed throughout the embedding, with cases falling neatly into regions corresponding to different conditions. This signals the potential for ProtAIDe-Dx to aid in the diagnosis of patients in early phases of impairment (formally tested in an external sample below). We also applied ProtAIDe-Dx to infer etiological diagnoses for the “Healthy AD” (diagnosis of AD but cognitive scores in healthy range), “Computed Dementia” (no diagnosis but cognitive scores in the dementia range), and Unknown (no diagnostic or cognitive information) groups (see Methods) and projected the resulting cases into the diagnostic embedding space (Fig. S3). Healthy AD and Computed Dementia cases were distributed along the control–AD axis, suggesting pathological heterogeneity, whereas most Unknown cases localized to the control region with a small subset in the stroke region.

There were many cases distributed on the embedding into regions inconsistent with their clinical diagnosis, i.e. AD cases distributed into stroke/TIA regions. This observation may have many interpretations – such cases could simply reflect failed model predictions, or even incorrect clinical diagnoses. However, these cases could also reflect cases with multiple overlapping pathologies or conditions with overlapping molecular etiologies or systemic presentations. Fig. 2F projects contours onto the embeddings representing the highest density of cases for each diagnosis, outlining the space in the embedding that individuals of each diagnosis typically occupy. In generating contours, we also noted sub-clusters of case densities distributed outside of the primary density. To investigate these trends, we clustered cases with the same diagnosis based on embedding coordinates (Fig. S4). Fig. 2F visualizes the top two clusters for each diagnosis.

The second (i.e. first non-dominant) cluster of healthy controls emerged at the intersection of shallow extremes of the AD and stroke/TIA regions. This group was older than the dominant cognitively unimpaired group (71.7±11.5 versus 57.7±15.8, FDR-corrected p=4e-56), worse educated, and had higher rates of hyperlipidemia (Fig. 2G, 28% versus 15%, FDR-corrected p=2e-6), other cardiovascular risk factors, more frequent history of smoking, higher cancer and diabetes rates and worse cognition (SuppData 2). Differential abundance analyses comparing the two control clusters revealed downregulation of proteins relating to inflammation in the dominant cluster compared to the minor cluster (SuppData 3, SuppData 4). These two clusters may resemble healthy (dominant) vs. unhealthy (minor) aging.

Two minor AD clusters also emerged, with one colocalizing in the stroke/TIA region, and the other in the PD region. Patients in these minor clusters showed lower rates of *APOE* ε4 carriage (FDR-corrected p=2e-7) and slightly better cognition (FDR-corrected p=3e-10). Patients in the minor cluster in the stroke/TIA region showed increased frequency of hypertension (Fig. 2H, 50% versus 26% and 35%, FDR-corrected p=0.01) and other vascular risk factors (SuppData 2), likely signaling mixed AD and vascular co-pathology, or misdiagnosis. Patients in the minor cluster within the PD region were younger and were predominantly male (69% vs. 43% and 46%, SuppData 2, FDR corrected p=0.03). Compared to the other clusters the dominant AD cluster had a higher abundance of proteins involved in cell death, damage response and mitochondrial activity, but lower abundance of proteins involved in immune and defense response (Fig 2J, Fig 2K). The minor cluster in the stroke/TIA region meanwhile had more abundance of proteins involved in immune response, plasticity and AD-related signals (i.e. APP and reelin processes) (SuppData 3, SuppData 4). Interestingly, both the minor clusters showed decreased abundance of proteins associated with energetic metabolisms, and in the case of the minor cluster in the PD region, specifically glucose metabolism (Fig. S5A, SuppData 3, SuppData 4). This is interesting given the robust finding of hypometabolism (especially relative to brain atrophy) in PD^56,57^, compared to AD, which can also produce phases of cortical hyperactivity^58^.

Perhaps most interestingly, a minor cluster for ALS emerged distant from the dominant cluster but in closer proximity to the FTD region. This minor cluster showed a higher rate of C9orf72 mutations (Fig. 2I, 50% versus 18%, FDR-corrected p=0.03), marginally higher rates of SOD1 mutations (Fig. S6, 50% versus 18.8%, unadjusted p=0.32), and a higher rate of mild cognitive impairment (SuppData 2, 99% versus 67%, FDR-corrected p=7e-11). Differential abundance between these two groups revealed upregulation of proteins relating to cell death in the C9orf72 cluster, and downregulation of proteins relating to metabolism and immunity (Fig. 2L, S5B, S5C; SuppData 3, SuppData 4). The dominant cluster meanwhile showed downregulation of proteins relating to muscle activity, tissue remodeling and neuroplasticity. Differential abundance and characteristics for PD, stroke/TIA and FTD can be found in SuppData 3, and SuppData 4.

**Figure 3.**
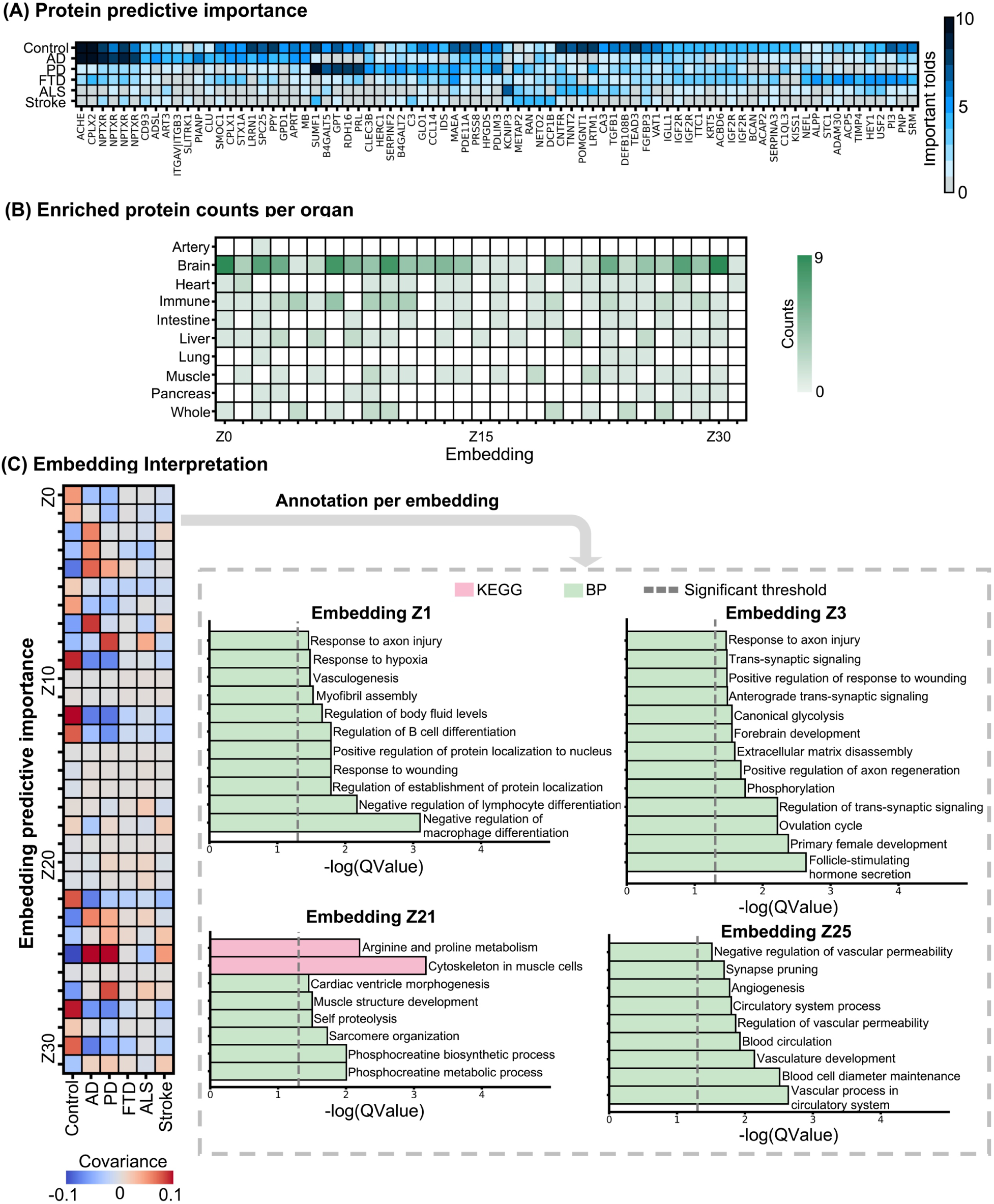
Model interpretations reveal proteomic content underlying model diagnostic predictions. (A) Predictive importance of proteins by fold counts. Proteins were visualized if they showed significant feature importance in more than 4 folds for at least one of the diagnostic tasks. (B) - (C) Interpretations for model-derived embeddings – nonlinear low-dimensional ensembles of proteins that contribute to predictions . (B) Frequency of organ-specific enrichment among embedding-specific proteins. (C) Left: Importance of embeddings for diagnostic tasks using covariance, which is a reliable feature importance metric for linear models proposed by Haufe et al., 2014; Right: Gene ontology enrichment analysis on selected embedding-specific proteins. Z1 shows signature for brain health or resilience; Z3 shows signature for AD; Z21shows signature for both ALS and PD; while Z25 may signal vascular dysfunction.

### Model interpretation highlights disease-specific networks and key discriminative proteins

While deep learning models are not trivial to interpret, understanding the underlying biological trends driving predictions made by ProtAIDe-Dx are essential for clinical adoption and biological insight. We employed a feature permutation approach at the inference stage^59^ to identify the most discriminative proteins used by our model. We employed a conservative approach, robust to multiple comparisons, highlighting proteins that were consistently associated with specific conditions across cross-validation folds (Fig. 3A, Fig S7). Several expected and previously described proteins emerged from this analysis, such as NEFL for FTD ^60–64^, CPLX^65,66^, CLU ^67,68^ and SMOC1 ^69–71^ for AD, and multiple NPTXR aptamers for multiple neurodegenerative diseases ^72,73^. ACHE was unsurprisingly an important discriminator for AD, given the common treatment of this condition with ACHE-inhibitors, and given the propensity of systemic medications to affect levels of circulating proteins ^74^. For PD, we replicated previous recent work^75^ highlighting SUMF1 as discriminative of PD, continuing the trend of lysosomal storage disorder-associated genes associated with PD^76^. SERPINF2 has also been reported as associated with PD by previous work^77^. Similarly, PRL and C3 have long been associated with PD, the former due to its relationship with dopaminergic function^78–80^, and the latter being indicative of immune changes detectable even before symptom onset^81–84^. Other interesting PD-discriminative proteins emerged. GPT has previously been shown to be discriminative in different synculeinopathies^85^, but related protein GPT2 has been implicated in midbrain neurodegeneration^86^. B4GALT-family proteins have been implicated in midbrain responses to environmental exposure^87^, while HERC1 is an endosomal system protein that has been previously linked to PD^88,89^.

A set of proteins was associated with both healthy controls and ALS, perhaps reflecting the model keying into the younger age or less prevalent cognitive impairment in these populations. For example, CNTFR is an astrocytic neurotrophic factor receptor that has been associated with better cognition and slower cognitive decline, in both CSF^90^ and plasma^91^, while TNNT2 is a dementia-associated heart-health protein^33,92,93^, which has been recently linked to ALS^94^. CNTFR, in particular, has a long history of interest as an ALS treatment due to its importance in the survival of motor neurons^95,96^. Meanwhile, POMGNT1 is likely more ALS-related, given its well documented association with congenital muscular dystrophy^97,98^, and LRTM1 plays a role in both brain and skeletomuscular systems^99^. KCNIP3 showed the strongest discriminative signal for ALS ^94,100,101^, though recent work has linked KCNIP3 expression to riluzole, a common treatment for ALS^94^. Certain proteins were discriminative of both ALS and FTD including HEY1, involved in Notch signaling and upregulated in mutant C9orf72 organoids^102^. However, several FTD-discriminating proteins captured our interest. SERPINA3 levels are altered in tissue of C9orf72 patients^103^, CSF levels differentiated symptomatic from unaffected FTD mutation-carrying patients^62^, and SERPINA3 levels were returned to normal after GRN-mutation mouse models were exposed to peripheral brain-penetrant GRN^104^. IGF2R has anti-inflammatory properties and is reduced across many neurodegenerative disorders, including in tissue of FTD patients and in GRN-insufficient cell models^105–108^. MAEA has been shown to interact with phosphorylated tau^109^ and TDP-43^110^, while CSF STC1 levels differ across neurodegenerative disorders^111^.

Looking at proteins discriminative of TIA/stroke, interestingly, several were also discriminative for AD (NPTXR, CD93) or PD (SUMF1, C3). While this may indicate that these proteins are useful for differentiating these diseases one from another, previous research instead suggests they may represent overlapping biological processes across disorders: neuronal pentraxin is a general signal of synaptic health^112^, while CD93 is likely involved in blood brain barrier integrity^113,114^, is altered in entorhinal endothelial cells in AD^115^, has been implicated in neurovascular disease^116,117^, and is generally a predictive marker of cognition^112^. C3 has also been reported as a meaningful marker in stroke patients^118–121^. Meanwhile, DCP1B has also been associated with neurovascular disease phenotypes^122^ and is discriminative for TIA/stroke in this study, but has previously been more associated with PD^123,124^, again possibly suggesting some shared pathways or etiologies among these disorders. Among those proteins most discriminative for TIA/stroke, METAP2 stood out as an angiogenesis regulator implicated in (among other things) obesity and stroke^125–127^, and RAN has been implicated in recovery from ischaemic events^128^.

Perhaps the most interesting proteins are those discriminative of healthy controls compared to other diseases, due to their potential as markers for general brain health. Many of the proteins associated with healthy controls seemed to be involved with discriminating them from other diseases, such as AD (e.g., ACHE, NPTXR, SMOC1, SPC25) or PD (e.g., SUMF1). However, some novel and interesting proteins were also found to be highly discriminative of certain conditions. For instance, GLO1 helps to manage mitochondrial and synaptic stress driven by advanced glycation end products, and has been shown to improve neurovascular coupling and bring about cognitive benefits in mice^129–131^. TGFB1 is critical to the transition between homeostatic and disease-responsive microglial states^132–134^, and while its levels in the brain have been associated with negative cognitive and neurodegenerative outcomes^135,136^, it may play an overall protective role^137^. VAT1^138,139^ and STX1A^140,141^ are both vesicular proteins that have been previously implicated in cognitive reserve, where the latter seems to be crucial for clearance of cellular debris via lysosomal exocytosis^142^. Both PDE11A^143–147^ and IGF2^108,148–150^ are expressed in limbic brain regions, play a role in memory processes, and have consistently been shown to rescue cognition in aging and neurodegeneration mouse models. PDE11A is a cAMP/cGMP-degrading enzyme that accumulates ectopically in the ventral hippocampus of both mice and humans with age^146,147^. IGF2, meanwhile, is neuronally enriched, involved in endosomal/lysosomal processes, is decreased in the hippocampus of AD patients and has also been shown to be neuroprotective in a PD context and various other contexts that involve abnormal protein accumulation^106,108,149^.

Given our interest in proteins discriminative of healthy cognition vs. all diseases, we ran a new model where SCD/MCI cases were now included as negative cases for all classes. This forced the model to distinguish cognitively healthy individuals additionally from mild (or in most cases, no objective) cognitive impairment or uncertain etiology, increasing specificity for cognitive health. Unsurprisingly, the model performed worse all around (Fig. S7), particularly for classifying recruited controls (77% vs 83%). However, the model interestingly identified OMG (oligodendrocyte myelin glycoprotein) as highly discriminative of cognitively unimpaired controls compared to cognitively impaired patients across all ten folds (Fig. S7). OMG binds to the NOGO receptor and inhibits neurite outgrowth, implicating it in axonal regeneration^151^. Large plasma proteomic surveys have found it to be less abundant in FTD patients^124^ and in patients with cerebral small vessel disease^152^, while lower CSF levels have been associated with β-amyloid levels^153^, confirming an association across multiple neurodegenerative diseases.

Another method for better understanding the ProtAIDe-Dx model is to probe the proteomic composition of the various embeddings that have been optimized toward these multiple diagnostic tasks. The embeddings should represent proteins that express unique nonlinear relationships in relation to neurodegenerative/neurological conditions, and therefore may represent isolated disease-relevant molecular networks or processes. We examine this underlying biology by observing the contribution of embeddings to diagnostic prediction, isolation and enrichment analysis of proteins involved in each embedding (Fig. 3C, SuppData 5, SuppData 6), and association between the embeddings and disease-specific biomarkers in an external dataset (Fig. 4C; see below). We employed a novel method for discovering the selected 648 proteins involved in each embedding (see Methods), though the approach does not indicate directionality of the effects. Contributing protein sets therefore represent a combination of co-abundant proteins and proteins with anti-correlated abundance within the embedding.

Given that proteins in blood come from organs throughout the body, we expected these embeddings to represent processes stemming from multiple organs. However, brain-specific proteins were highly prevalent across all embeddings (Fig. 3B). We therefore tested for enrichment of specific neural cell types (SuppData 7, SuppData 8). Embedding Z2 showed strong specificity to the brain, and to neurons in particular (SuppData 7). Z2 was also discriminative of cognitively unimpaired, AD, and stroke/TIA diagnoses (Fig. 3C), and enrichment terms relating to neuronal resilience, synaptic communication, and molecular homeostasis (SuppData 5), and was associated with older age, worse cognition and unhealthier levels across multiple markers in an external sample (Fig. 4C). This embedding may represent neuronal functional decline, reflecting reduced resilience and synaptic dysregulation that contribute to cognitive impairment across aging and neurodegeneration. Embedding Z23 differentiated controls from neurodegenerative conditions and was enriched for proteins expressed by oligodendrocyte precursor cells (OPC, SuppData 7, SuppData 8). The associated enrichment terms included immune activation, structural remodeling, cytoskeletal organization, proteostasis, vascular function, and developmental processes (SuppData 5). This embedding was also associated with older age, adverse biomarker profiles, and male sex. Together, these features suggest that Z23 may capture glial vulnerability pathways that link aging and sex to increased neurodegenerative disease risk.

Other embeddings emerged with interpretable annotations helping to understand proteomic underpinnings of specific neurodegenerative diagnosis (Fig. 3C, SuppData 5, SuppData 6). Embedding Z3 showed specificity for AD (Fig. 3C) and was associated with worse cognition and AD biomarkers in an external sample (Fig. 4C). The enrichment in sex-related reproductive and hormonal regulation suggests that part of the signal may reflect female sex differences in AD risk, a rapidly expanding line of research^154^. In addition, enrichment for OPC proteins, synaptic activity and remodeling, phosphorylation, and tissue damage (Fig. 3C, SuppData 7, SuppData 8) may reflect processes linking sex-related pathology with cellular vulnerability, synaptic dysfunction, and injury in AD. Embedding Z21 was discriminative for both ALS and PD diagnosis and was associated with younger age, better cognition, and healthier levels across multiple markers in an external sample (Fig. 4C). Z21 showed specificity to both brain and muscle systems (Fig. 3B) and was enriched for proteins involved in muscle energy buffering, contractile apparatus integrity, and structural remodeling, which are critical for muscle performance and therefore highly relevant to motor and movement disorders. Embedding Z25 was discriminative for both AD and PD diagnosis and represented the most important embedding for stroke/TIA classification (Fig. 3C). In an external sample, Z25 was associated with older age, poorer cognition, and biomarkers of vascular pathology, neurodegeneration, and inflammation (Fig. 4C). Functional enrichment analysis indicated that Z25 was linked to proteins involved in vasculature development, angiogenesis, blood circulation, and the regulation of vessel diameter and permeability, suggesting a signal of vascular activity and potential blood–brain barrier permeability. Finally, some embeddings appeared to capture proteomic features associated with brain health or resilience (Fig. 3C, SuppData 5, SuppData 6). For example, embedding Z1 discriminated cognitively unimpaired individuals from patients (Fig. 3C). In an external cohort, it was associated with tau pathology, but otherwise with better cognitive performance, and generally healthier biomarker profiles (Fig. 4C). The enriched terms of Z1 underscore processes related to responses to damage and hypoxia, protein response to the nucleus, vascular and metabolic homeostasis, and negative regulation of inflammatory and complement systems. Together, these findings suggest that embedding Z1 may capture protective or resilience signals, or may be capturing a very early disease response. A more comprehensive view of shared GO terms between embeddings and biological similarities of embeddings confirmed by biomarkers could be found in Fig. S8.

**Figure 4.**
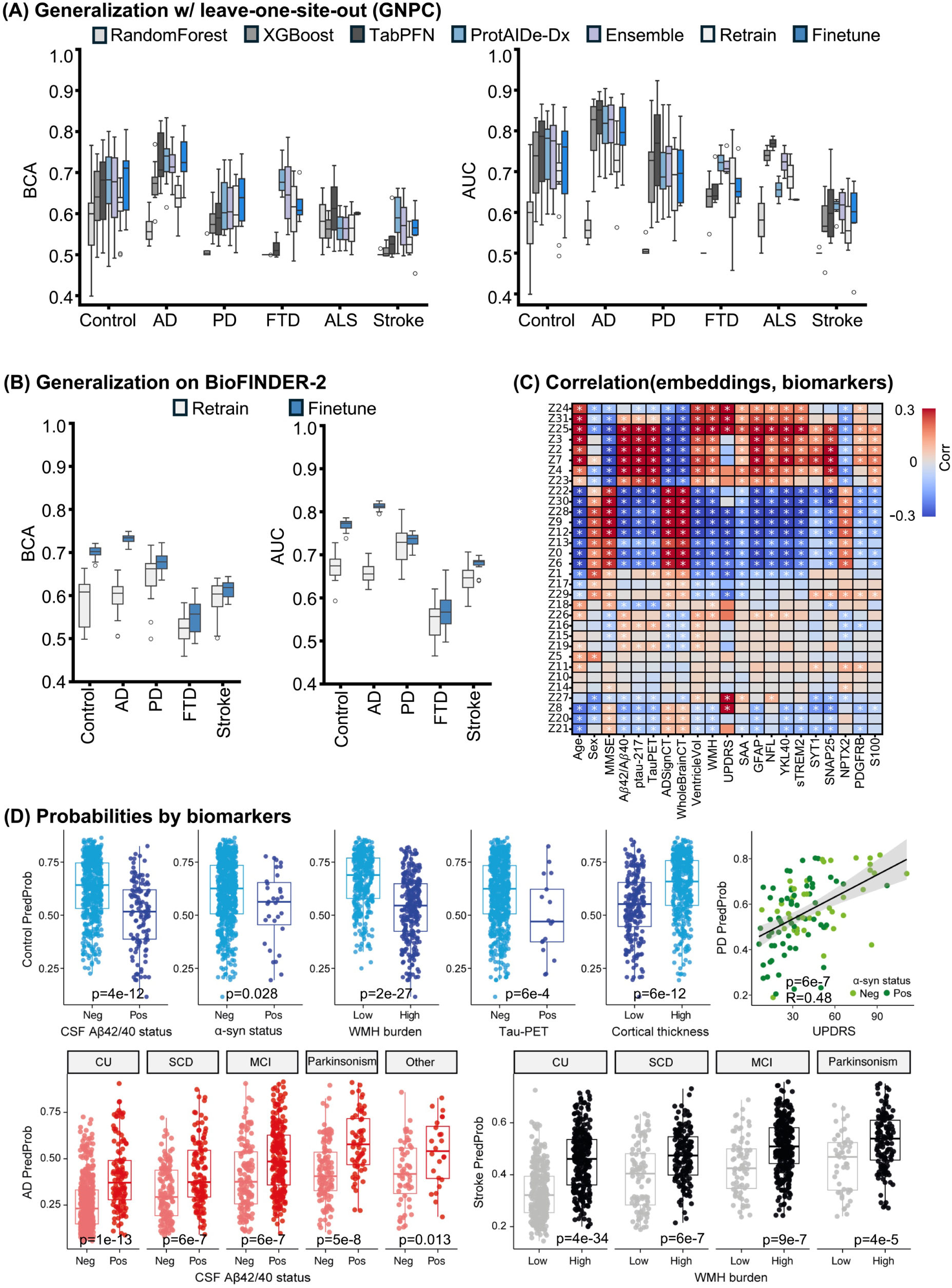
Model validation on external BioFIDNER-2 cohort. (A) Leave-one-site-out balanced classification accuracy (BCAs; left) and AUCs (right) of ProtAIDe-Dx on GNPC. Retrain: Retrain logistic regression model on K=100 participants’ proteins, FineTune: Retrain logistic regression model on K=100 participants’ proteomics embeddings. (B) Model generalization performance on BioFINDER-2 cohort. Left: BCA; Right: AUC. (C) Correlation between embedding and biomarkers, white star indicates a significant relationship after FDR correction across all comparisons. (D) Diagnostic predictive probabilities for control (blue), AD (red), parkinsonism (PD; green) and stroke/TIA (black) diagnoses, associated with different biomarkers, stratified by biomarker-confirmed clinical diagnosis. Independent t-tests were used to compare group differences, and p values were corrected for multiple comparisons using FDR.

### Out-of-sample generalization and validation by biomarkers of disease-specific neuropathology

A possible limitation of ProtAIDe-Dx is that it is trained exclusively on clinical diagnosis, and most of these diagnoses were performed without biomarker support. Therefore, we sought to validate ProtAIDe-Dx in an external dataset with many disease-specific biomarkers (BioFINDER-2 dataset, N=1,786). Out-of-sample generalization is necessary for AI models such as ProtAIDe-Dx to be translated to clinical settings, but is challenging given that within-sample performance tends to be optimistic. In our case, this issue is likely exacerbated by high variation in effect sizes of individual proteins across sites (Fig. S9). We therefore used a leave-one-site-out cross-validation approach to evaluate the performance of ProtAIDe-Dx in out-of-sample generalization tasks. ProtAIDe-Dx continued to outperform the Random Forest and XGBoost baselines significantly and showed slight improvements over TabPFN in FTD and stroke prediction, but generally took a substantial dip in performance in both balanced accuracy score and AUC compared to whole-sample cross-validation performance (Fig. 4A, Table S4, SuppData 9). Unlike in the whole-sample cross-validation analysis, this time, the ensemble model did not aid performance. However, model performance was partially recovered by using finetuning (see Methods), a process involving calibration of a model to a new dataset using a select number of exemplary cases. This approach was particularly effective for improving discrimination of PD patients. Additional site-to-site generalization experiments revealed that there was around ∼10% drop in performance when generalizing across sites (Table S5, Fig. S10).

When applying ProtAIDe-Dx to the BioFINDER-2 dataset, diagnostic performance was close to the median of general leave-one-site-out performance (Fig. 4B). Looking at predicted probabilities across diagnosis revealed some expected trends (Fig. S11). For example, PD probabilities were elevated in PD patients, but also in dementia with Lewy bodies (DLB) cases, and stroke/TIA probability was elevated in patients with vascular dementia. Subsequently, we were interested whether these probabilities correlated with disease-specific biomarkers within disease groups (Fig. 4D, S12). Among cognitively unimpaired individuals, the model-derived probability of being cognitively unimpaired was lower for participants expressing AD (FDR-corrected p < 0.05 for all AD biomarkers), Lewy body (FDR-corrected p = 0.028 for CSF ɑ-synuclein) or neurovascular pathology (FDR-corrected p = 2e-27 for WMH) (Fig. 4D). This nominates cognitively unimpaired probabilities as a potential signal of general neurodegenerative malaise, and indicates that some “false positive” results from ProtAIDe-Dx are actually correctly identifying underlying preclinical neuropathology (Table S6). Similarly, AD probabilities were higher in non-AD cases with comorbid Aβ and Tau pathology (Fig. 4D, S13), and higher stroke/TIA probabilities were associated with greater WMH burden in both impaired and unimpaired individuals (Fig. 4D). PD probabilities did not show a significant relationship with presence of Lewy body pathology (as measured using CSF ɑ-synuclein seed amplification assays; SAA), but were correlated with symptom progression (UPDRS) in PD cases (R=0.48, Fig. 4D). This latter finding is important given a lack of continuous in vivo biological signatures of PD progression.

**Figure 5.**
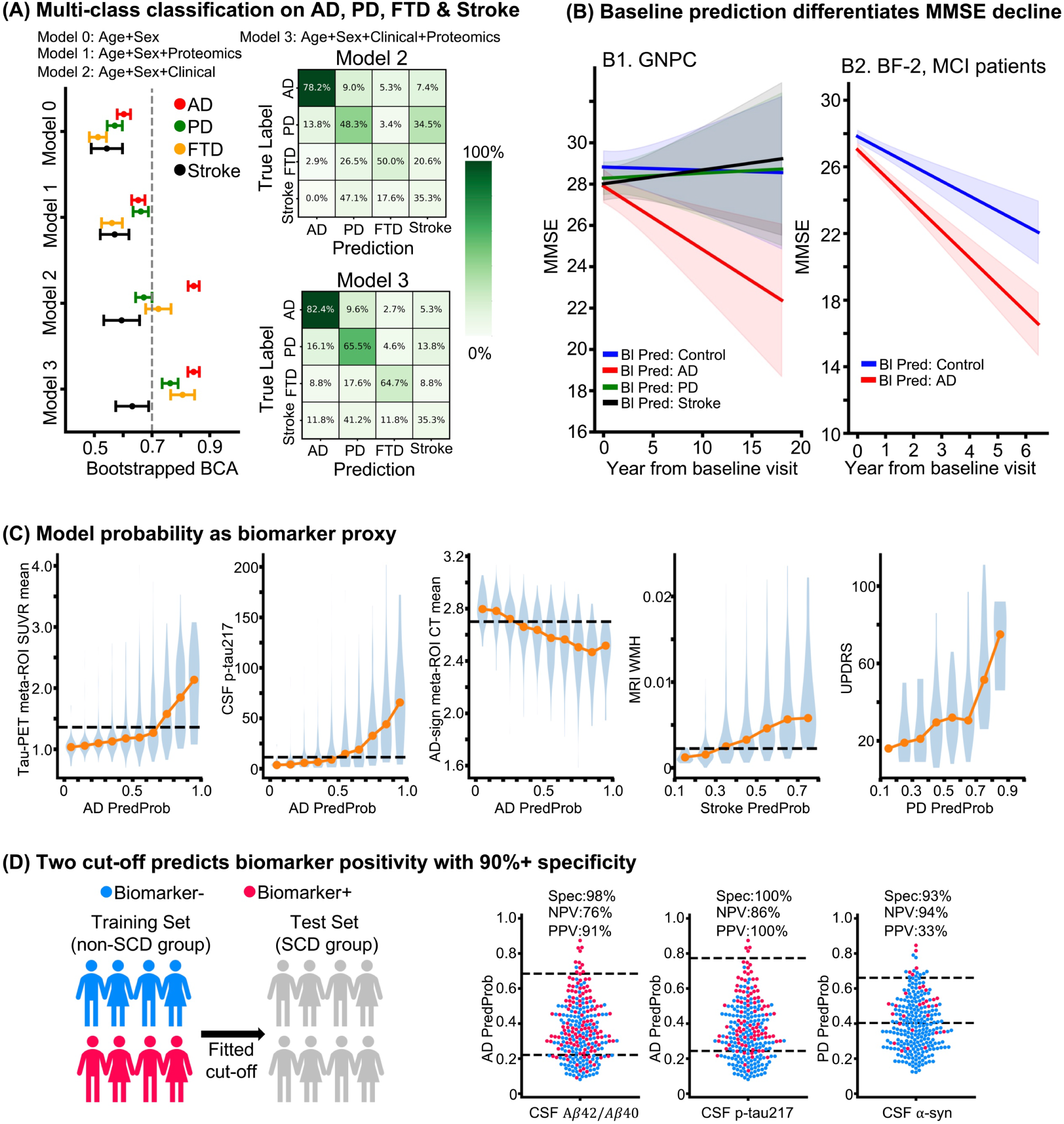
Clinical Utility of ProtAIDe-Dx. (A) Multi-class classification on AD, PD, FTD, and Stroke patients using various models inclusive of demographics, ProtAIDe-Dx embeddings, and/or common clinical biomarkers. Left: One-versus-rest balanced classification accuracy (BCA) computed by 1000 bootstraps on testing patients, Right: Confusion matrix on testing patients for Model 2 (top) and Model 3 (bottom). (B) Left: Model-predicted baseline diagnoses differentiated longitudinal MMSE trajectories of GNPC patients irrespective of true baseline clinical diagnosis. Trajectories were modeled using linear mixed effect model “MMSE ∼ Age + Sex + Site + BaselineDx + BaselinePred * Year + Year | SubjectID”. Right: Replication on BioFINDER-2 MCI patients. (C) Biomarker distributions across predicted probability bins. SUVR, Standardized Uptake Value Ratio; CT, cortical thickness; WHM, white matter hyperintensity; UPDRS, Unified Parkinson’s Disease Rating Scale. (D) Two-cutoff strategies for predicting biomarker positivity. Cutoffs were derived from non-SCD BioFINDER-2 participants to achieve 90% NPVs and PPVs, except for α-synuclein, where the PPV was set at 40% owing to sample validity constraints. These fitted cutoffs were then applied to SCD patients to estimate accuracy in this clinically relevant sample.

### Proteomics provide additive information to diagnosis in a memory clinic sample

For models like ProtAIDe-Dx to be translated to real-life clinical applications, it is important to show evidence that they provide additive value in these settings. Therefore, using the same external BioFINDER-2 dataset (not involved in model training), we generalized ProtAIDe-Dx to a differential diagnostic task in a memory clinic sample (i.e., patients with subjective or objective cognitive impairment seeking memory clinic care). Due to advanced biomarkers, we have greater insight into the probable primary etiological diagnosis of AD, PD, FTD or Stroke/Vascular dementia in these cases, and we fit a series of models seeking to identify this primary diagnosis using a baseline model of only age and sex (Model 0), using just ProtAIDe-Dx and demographics (Model 1), using accessible clinical markers (Model 2: demographics, MMSE, mean cortical thickness of AD-signature meta-ROI (ADSignCT, ^155^), plasma p-tau217, and plasma NEFL), and using all of these markers together (Model 3). The final model incorporating ProtAIDe-Dx with common clinical biomarkers (Model 3) achieved significantly better balanced classification accuracy than the model using only common clinical markers (Model 3), especially adding value in diagnosis of non-AD dementias (Fig. 5A, Table S7, S8). This shows that ProtAIDe-Dx embeddings provide information complementary to existing clinically accessible neurodegenerative disease biomarkers. To test the limits of ProtAIDe-Dx’s capability for extension to novel differential diagnosis tasks, we tested whether ProtAIDe-Dx embeddings could distinguish PD (n=78), DLB (n=48), PSP (n=34), and MSA (n=16) in the BioFINDER-2 cohort. This task resulted in an accuracy of 0.52 ± 0.04, which was significantly greater than the null accuracy of 0.44 (p = 0.012), notable given that ProtAIDe-Dx was not trained for this specific task.

Although ProtAIDe-Dx was trained exclusively on baseline visits, it demonstrated the ability to differentiate longitudinal rates of cognitive decline. We fitted linear mixed-effects models, with individuals as random effects, to assess whether baseline characteristics (e.g., clinical diagnoses, model predictions) could predict longitudinal trajectories of decline. Compared with baseline clinical diagnoses, which did not distinguish rates of decline after FDR correction in the GNPC (p > 0.05 across all diagnostic groups; Fig. S14, Table S9), baseline predicted diagnosis from ProtAIDe-Dx significantly stratified decline trajectories (FDR-corrected p < 0.05 across all prediction groups; Fig. 5B1, Table S10), independent of clinical diagnosis. Specifically, individuals predicted as AD exhibited faster cognitive decline than those predicted as Control, PD, or stroke. These findings were replicated in the external BioFINDER-2 dataset, where MCI patients predicted as AD by ProtAIDe-Dx declined more rapidly than MCI patients predicted as Control (FDR-corrected p = 0.0015; Fig. 5B2, Table S11).

The probability outputs from ProtAIDe-Dx provide clinically interpretable indicators of biomarker status. In the external BioFINDER-2 dataset, we observed strong correlations between predicted probabilities and corresponding biomarker measurements (Fig. 5C). Specifically, when the AD probability exceeded 0.9, most patients were tau-positive by TauPET (first column) or CSF p-tau217 (second column) and showed cortical thickness below the diagnostic threshold (third column). Likewise, when the stroke probability exceeded 0.7, most patients exhibited elevated white matter hyperintensities (fourth column). Motivated by these strong correlations, we implemented a two cut-off strategy to classify biomarker positivity versus negativity. The cut-offs were derived from non-SCD participants in the external BioFINDER-2 dataset and subsequently tested on SCD patients. This approach achieved >90% specificity for detecting CSF-based biomarker positivity. For AD-related biomarkers, the method yielded 90% PPV for both CSF amyloid-β and p-tau217. In addition, it achieved 94% NPV for detecting LBD-related biomarker positivity based on CSF α-synuclein. We also show results of models optimized on PPV/NPV, and models ensuring 50% coverage (Fig. S15).

**Figure 6.**
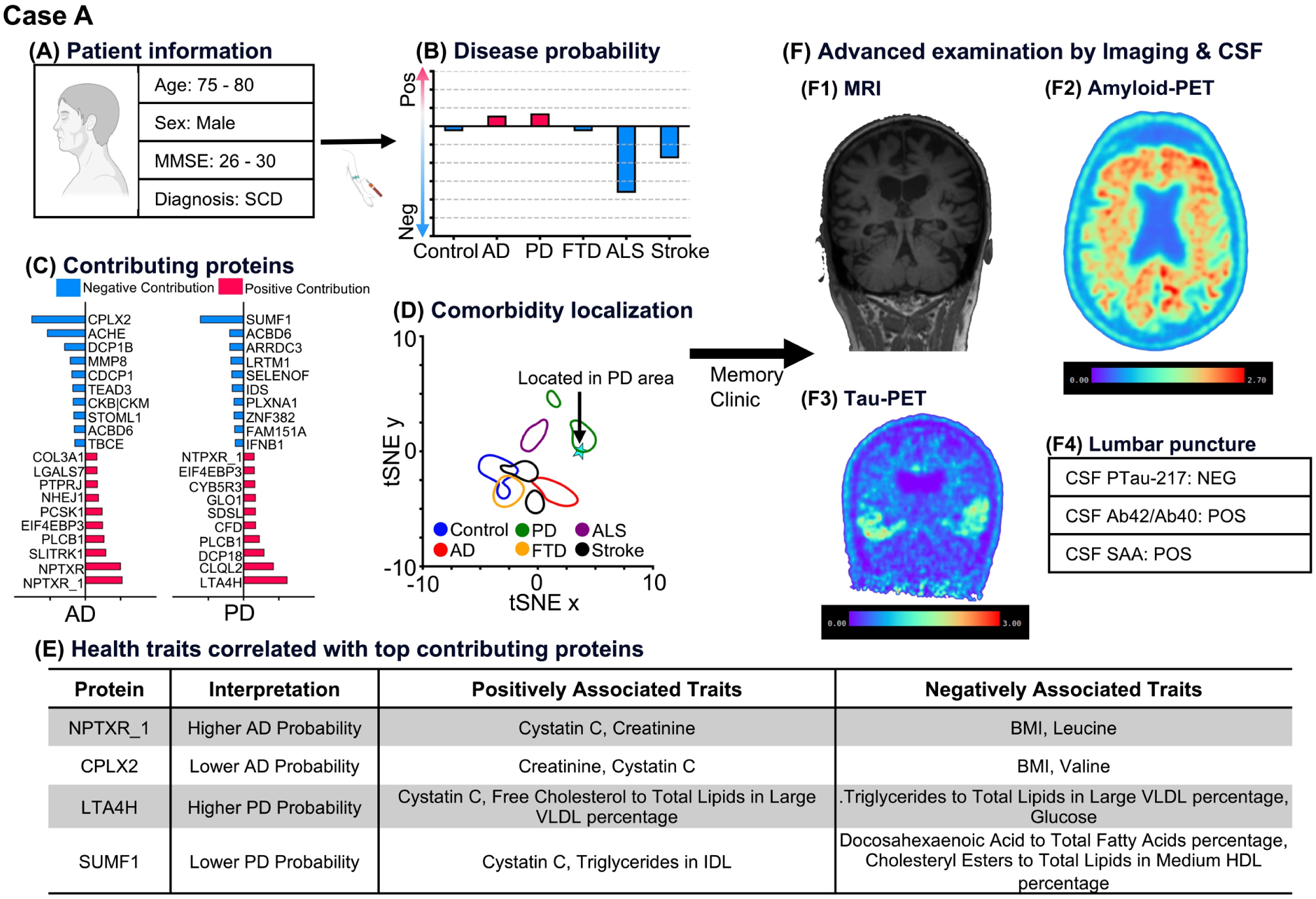
Individual neurodegeneration risk report (Case A). (A) Demographics and cognition score of one older participant, who was diagnosed with SCD. (B) ProtAIDe-Dx predicted this participant with higher probabilities of AD and PD. The probabilities were normalized to be centered at zero. (C) Contributing proteins for making the decision were computed based on SHAP values. The top 10 positive and top 10 negative proteins were visualized. (D) The location of this participant on the diagnostic probability map indicates his proteomic signature was consistent with typical PD patients. (E) Health traits correlated with top contributing proteins were listed, informing clinicians to pay attention to these traits. (F) A signature such as this would be suggestive of follow-up advanced imaging and CSF biomarker examination at a memory clinic to confirm diagnosis. Here, AD pathology was confirmed by both a positive β-amyloid-PET scan and a positive tau-PET, as well as a positive CSF Ab42/40 ratio, but not positive CSF p-tau217 levels, interestingly). Presence of underlying Lewy body pathology was confirmed through a positive CSF seed-amplification assay for ɑ-synuclein pathology.

### Proof-of-concept diagnostic report by ProtAIDe-Dx

The clinical utility analysis shows the potential of ProtAIDe-Dx to provide significant additive information to clinical diagnostic work-up. We therefore built a proof-of-concept for a diagnostic report using ProtAIDe-Dx (Fig. 6, S16, S17). The report indicates diagnostic probability across all conditions (Fig. 6B), and includes a localization of the patient to the GNPC disease probability space (Fig. 2A; Fig. 6D). Importantly, the report uses model explanation technology to report which proteins contributed to this specific individual’s prediction (Fig. 6C), allowing a clear biological explanation of the proteomic diagnosis. Physical traits linked to these patient-specific diagnostic proteins are also provided by programmatically accessing a library of protein-trait associations (Fig. 6E), providing possibility for lifestyle interventions or further explanation.

We use three patient case examples to showcase the report. Case A (Fig. 5) was a 75-80 year-old male entering the memory clinic with subjective cognitive complaints but objectively intact cognition. ProtAIDe-Dx predicted underlying comorbid AD and Lewy body pathologies. This was confirmed by PET scans showing cortical Aβ burden and temporal tau pathology (AD), as well as positive CSF SAA indicating Lewy body pathology (Fig. 5F). Case B (Fig. S16) presented with MCI in their late-60s, and was given a probable diagnosis of AD by ProtAIDe-Dx, likely due to proteins indicating AD-related Aβ42 (PLCB1), and neuroinflammation (CFD). CSF and PET biomarkers confirmed the presence of AD Aβ and tau pathology in this patient. Finally, Case C was a late-70s male without any subjective or objective cognitive impairment, recruited from the Malmö population. ProtAIDe-Dx predicted a diagnosis of cognitively unimpaired with underlying neurovascular disease (Fig. S17). CSF and PET biomarkers were negative for AD pathology, but the MRI showed enlarged lateral ventricles, periventricular WMH burden and widespread cortical atrophy indicative of neurovascular disease. While not all ProtAIDe-Dx probabilities were accurate and confirmed by biomarkers (Fig. 4B,E), these examples showcase the potential of ProtAIDe-Dx in adding explainable proteomic data informative toward neurodegenerative etiology to clinical workup.

## Discussion

The rapid growth of dementia populations worldwide urges scalable and accessible biomarkers for neurodegenerative diseases. Such biomarkers will undoubtedly revolutionize various fields of biomedicine, including pharmaceutical (e.g., trial recruitment, ^156^), clinical (e.g., early-stage intervention, ^157^), and research (e.g., pathology, ^158^) pursuits. Following the recent success of AD biomarkers, biomarkers for many other neurodegenerative diseases are in development, such as extracellular vesicle (EV) biomarkers for FTD pathologies ^159^ and CSF seed amplification assays of ɑ-synuclein for Lewy Body diseases ^160^. However, while promising biomarkers for certain diseases are on the horizon ^161^, many biomarkers under development are either invasive (CSF) or not yet scalable, and are usually singular, requiring multiple tests to assess different pathologies. Therefore, a one-shot, multi-disease biomarker panel that is minimally invasive, economical, and widely accessible is highly anticipated. This study presents an early attempt to apply deep neural networks on a massive (N=17,187) neurodegenerative disease sample and high-rank (7,595 proteins) plasma proteomics data, toward this goal. Our proposed ProtAIDe-Dx model synthesized novel biological insight and exhibited clinical utility by mining neurodegeneration-related signals from high-rank proteomics data at multiple levels. These levels ranged from discovering key proteins for robust prediction of certain diseases, to extraction of neurologically relevant protein networks, to simultaneous generation of probabilistic diagnoses across several neurological conditions. Moreover, when generalized to an out-of-sample memory clinic dataset, ProtAIDe-Dx improved automated differential diagnosis accuracy beyond the capability of currently accessible clinical biomarkers. This study distinguishes itself from other neurodegenerative disease proteomics studies ^162–165^ due to the focus here on making individual-level predictions on new cases, by training a model to recognize multiple co-pathologies rather than on differential diagnosis, and by rigorously avoiding leakage and overfitting and providing true generalization performance. However, despite providing complementary information capable of enhancing current clinical workup, our data suggests that contemporary high-throughput plasma proteomics assays alone cannot yet replace currently available clinical markers. Altogether, this study sets a benchmark for future proteomics studies by providing a baseline for predictive performance, a resource sorely needed in the biomedicine AI field ^166^. Below, we highlight promising findings, and we outline key challenges to be addressed in future work.

The performance of ProtAIDe-Dx is currently not sufficient to replace currently available clinical markers, though we demonstrate several that it could be integrated into clinical practices as an accessible and cost-effective assistant. Disease-differentiation experiments indicated that ProtAIDe-Dx provides additive value beyond currently available clinical biomarkers, highlighting its clinical potential given the high prevalence of neurodegenerative co-pathologies in the aging population and the fact that affordable, minimally invasive biomarkers for PD and FTD are still under development ^159,160^. ProtAIDe-Dx’s predictions to differentiate longitudinal rates of cognitive decline may offer a useful tool for clinical management, as individuals predicted as AD tended to exhibit faster decline irrespective of their current diagnostic status. These findings suggest that diagnostic labels may not fully reflect underlying disease progression, whereas ProtAIDe-Dx tends to capture more informative biological signals that could enhance clinical decision-making. Such “predicted AD” individuals could therefore warrant closer monitoring and may benefit from consideration of earlier intervention strategies to potentially delay progression to dementia. Probability outputs from ProtAIDe-Dx provide clinically interpretable indicators of biomarker status, consistent with established approaches such as the Amyloid Probability Score 2 (APS2) developed by C2N Diagnostics. Higher predicted probabilities generally reflected greater biomarker burden, and the two cut-off approach achieved >90% specificity for determining biomarker positivity. In clinical settings without access to advanced biomarker or imaging assessments, such probabilistic estimates from ProtAIDe-Dx could serve as an initial proxy, potentially improving diagnostic efficiency and accuracy. In more well-resourced scenarios, ProtAIDe-Dx can provide complementary information on disease-associated proteomic changes that can help inform more confident diagnoses.

Due to its diverse site composition and unstandardized data collection, the GNPC creates an excellent dataset for building and testing tools for translation to real-world clinical settings, where data curation and standardization rarely match that of most research datasets. Within the GNPC, ProtAIDe-Dx achieved impressive cross-validated diagnostic accuracy across diseases, especially for ALS and PD. However, this performance decreased in a leave-one-site-out validation setting and a site-to-site generalization setting that more closely simulates a true clinical translation, especially for disease with high imbalanced distribution across sites (e.g., ALS patients mainly come from one site). This indicates that, despite the large and diverse sample, site effects and overfitting remain a challenge for training generalizable models ^167,168^. This cannot necessarily be explained by over-complexity of our deep learning architecture, as ProtAIDe-Dx consistently outperformed the less complex random forest and XGBoost models. A similar performance drop was observed in the state-of-the-art TabPFN model, which was trained on millions of synthetic datasets, indicating that existing site effects cannot be overcome solely by simply increasing sample size. It is likely that advances in generalizable data harmonization ^169,170^ or standardization ^171^ will be necessary to overcome these limitations. Model finetuning ^172,173^, a realistic application for a real out-of-site translation scenario, did help to recover performance, particularly for PD.

In general, several factors must be considered when evaluating the diagnostic performance of ProtAIDe-Dx. One of these factors is the difficulty of the task. For instance, FTD is a highly heterogeneous disorder with multiple underlying pathologies and many different clinical presentations ^174,175^. Our model was trained on only 206 FTD participants but, despite these limitations, still managed greater-than-chance performance in out-of-site data, and outperformed all baseline and even state-of-the-art models. This suggests that there are biological features consistent among FTD patients but unique to FTD disorders, which could likely be exploited with larger samples and more in-depth annotation of patients. Similarly, we used stroke/TIA as a proxy for confirmed neurovascular disease. Prediction of stroke/TIA patients is a challenging task, given that it is a stochastic event that individuals with neurovascular disease are at risk for, rather than a disease itself. Furthermore, stroke is only one among many manifestations of neurovascular disease ^35,176^, and itself may often go undiagnosed. Despite these challenges, ProtAIDe-Dx learned a plasma proteomic pattern that not only provided greater-than-chance prediction of stroke patients, but also generalized to other indicators of neurovascular disease in a separate cohort.

However, one of the most important considerations in evaluating the performance of prediction models in neurological diseases relates to the accuracy of the original clinical diagnosis, which both forms the basis of training and serves as the standard for evaluation. Neurological and neurodegenerative diseases are notoriously difficult to diagnose ^177,178^, and in our particular case, models were trained and evaluated based on clinical diagnoses that, for the most part, lacked biomarker confirmation. In addition, the clinical diagnosis criteria varied across sites, given that GNPC is a retrospective collection of multiple cohorts. Given that there are multiple criteria for the diagnosis of AD ^179–181^ and multiple interpretations of those criteria, this issue will be present in any real-world application. In light of these limitations, many “false” predictions from ProtAIDe-Dx may not represent incorrect diagnoses, considering the prevalence of asymptomatic disease, co-pathology, and misdiagnosis. These concerns were validated on an external dataset with biomarker information. For example, we found that the model tends to predict cognitively unimpaired participants with older age, higher rates of abnormal CSF Aβ42/Aβ40 ratio, and higher CSF p-tau217 as AD dementia.

Along similar lines, in the main GNPC dataset, we found clusters of patients with one diagnosis that showed proteomic signatures in their blood plasma indicative of different diagnosis or etiology. For example, we found a minor cluster of AD patients with proteomic patterns similar to stroke/TIA patients. This might indicate misdiagnosis, could be indicative of co-pathology ^182^, or may even indicate a vascular etiology for a subgroup of AD patients ^183,184^. Supporting the latter hypothesis, Tijms et al. described a proteomic AD subtype associated with blood-brain barrier dysfunction ^164^ that exhibited similar molecular features to our vascular-AD cluster, including increased levels of immunity-related proteins and reduced synaptic-related processes. By contrast, the “dominant” AD cluster in our study was characterized by a higher abundance of proteins involved in metabolism, cell death, and stress response, alongside lower levels of immune and defense-related proteins. These molecular patterns are corroborated by previous studies. Johnson et al. identified several modules related to AD, including one enriched for MAPK signaling and metabolic pathways, where MAPK is known to regulate cell survival, cell death, and stress responses, and another specifically associated with sugar metabolism ^185^. Dysfunction in energy metabolism and brain immunity has also been recognized as a key alteration in AD pathophysiology in another study, with increased metabolic and glycolytic activity and decreased immune-related proteins emerging later in the disease continuum ^34^.

As another example of proteomic signatures perhaps signaling differing underlying etiologies, we also observed a sub-cluster of ALS patients with a higher rate of C9orf72 mutation carriers, that exhibited a proteomic signature more similar to FTD patients. This highlights the similar underlying molecular pathways known to be shared by some ALS and FTD patients ^186^. Interestingly, differential abundance analysis revealed an upregulation of proteins related to cell death in this C9orf72 ALS cluster. Activation of programmed cell death, including apoptosis and other regulated death pathways, is a well-established feature of ALS pathology, observed in both sporadic and familial cases as well as in ALS mouse models ^187^. Interestingly, C9orf72 has been implicated in the activation of p53, a major upstream regulator of apoptosis, with evidence showing that mutant C9orf72 promotes neurodegeneration through p53-driven cell death ^188^. This raises the possibility that p53-related mechanisms may represent a cell death pathway more specifically engaged in C9orf72-associated ALS.

Findings of clinical syndromes demonstrating distinct proteomic profiles are in line with a growing number of studies finding multiple proteomic or transcriptomic subtypes within the AD population ^164,189–191^. Together, these observations demonstrate that there is a set of neurodegenerative signatures detectable in blood plasma that may be associated with differing clinical profiles, or that may represent stratified responses to general neurological insult. In either case, a patient’s underlying biological response to pathology may be just as relevant to treatment and prognosis as their clinical presentation. Future work should leverage longitudinal data to further explore the clinical meaningfulness of these proteomic disease subgroups.

We saw many cases where ProtAIDe-Dx seemed to be learning a signature associated with a specific neuropathology, rather than with a clinical syndrome per se. However, we also observed scenarios where ProtAIDe-Dx appeared to be more attuned to symptoms than to underlying biology. When generalized to a new sample, our model’s PD probabilities were not associated with CSF ɑ-synuclein status, but were instead correlated with symptom progression in PD cases. While somewhat disappointing with respect to differential diagnosis of motor disorders, there is still clinical potential here, given that no continuous biological measures for tracking PD progression are currently available. On a similar note, we found that lower control probabilities were associated with the presence of multiple pathologies in the cognitively unimpaired group, underscoring the potential of this signature as a measure of general brain health^192^.

Beyond producing diagnostic probabilities for multiple diseases, ProtAIDe-Dx also identified a compact set of top predictive proteins that significantly influenced model performance on out-of-sample data. Many previous studies have described lists of proteins associated with different diseases ^30,34,35,49,185,193,194^, and it is important to distinguish between predictive and associational findings. With larger sample sizes and a richer set of variables, association analyses are more prone to detecting “significant” spurious and indirect associations between variables and group status. In contrast, predictive modeling provides a more reliable assessment of a variable’s discriminative roles, given that it is validated on out-of-sample data ^195^. As a result, predictive findings tend to be more conservative and potentially more clinically useful. In our study, among the top proteins discriminating different conditions, those proteins that discriminated controls from all neurodegenerative conditions were perhaps the most interesting. GLO1, PDE11A and IGF2 have all been investigated as cognition enhancers in various aging models^130,131,147–149^ and may indeed be promising targets given that they also signal healthy aging in our large human cohort. Many of the proteins profiled were brain expressed and played a role in either memory (PDE11A, IGF2), inflammation (TGFB1), neurite regeneration (OMG) or endosomal/lysosomal systems (STX1A). OMG, was a highly discriminative protein for controls specifically when differentiating them from not just dementia but also MCI-SCI patients, suggesting its possible role in maintenance of cognition. Meanwhile, lower expression levels of NPTXR in neurodegenerative groups have been observed in multiple studies (^73,112,196^, highlighting synapse loss as a common feature of neurodegeneration ^72^. These proteins could, in combination, form the basis of a low-cost blood test for multi-cause neurological malaise, which might be useful in triaging or early diagnosis and monitoring. Further studies should continue to investigate how these proteins change over time in response to other outcomes of brain health.

Some key proteins for neurodegenerative diseases were identified by ProtAIDe-Dx as well. NEFL, involved in neuronal damage, has been commonly found to have increased concentrations and in multiple neurodegenerative diseases ^197–199^, including FTD ^36,200^, making it unsurprising as a protein predictive of FTD in the present study. Interestingly, PI3 was found to be discriminative for FTD. PI3 has previously been reported to be abundant in the plasma of cognitive impairment and dementia groups^201^ and predictive for future dementia risk^47^. Given that PI3 encodes elafin, a serine protease inhibitor with potent anti-inflammatory activity ^202^, and that all major genetic forms of FTD exhibit elevated inflammatory signatures^203,204^, PI3 may influence FTD pathology by modulating neuroinflammatory responses, either as a compensatory protective factor or as a biomarker of inflammatory dysregulation, a possibility that warrants further experimental validation.

ProtAIDe-Dx also identified several proteins predictive of stroke/TIA, perhaps the most promising being RAN. RAN, traditionally recognized for its roles in cell division and signaling ^205,206^, has recently emerged as a modulator of oxidative stress, apoptosis, and glial activation in models of ischemic brain injury^128^. By conferring neuroprotection and attenuating neuroinflammation, it represents a promising therapeutic target for stroke and related neurovascular disorders, further supported by genetic evidence linking an RNA variant (rs1435) to post-stroke mortality in patients with large-artery disease^207^. Another interesting protein predictive of stroke/TIA was MATAP2. MATAP2, best known for its role in angiogenesis ^208^, has also been implicated in stroke. Experimental ischemia shows calpain-mediated cleavage of MATAP2, linking it to stress pathways during neuronal injury^126^, while its functions in endothelial remodeling^209^ and glial inflammation^210^ suggest it may influence both acute injury and recovery. Together, these findings highlight MetAP2 as a potential but underexplored therapeutic target in neurovascular disease.

KCNIP3 was identified by ProtAIDe-Dx as predictive for ALS, in line with previous studies suggesting its involvement in the disease. It has been listed among ∼300 ALS-associated genes^211^ and reported to be upregulated in ALS patients^212,213^, with similar observations in animal models^214^. In plasma proteomics, KCNIP3 levels were found to be increased by approximately 60% in ALS patients receiving riluzole treatment^94^. Functionally, KCNIP3 binds voltage-gated potassium channels and regulates A-type currents, which counteract depolarizing inputs and modulate neuronal excitability^215^. These findings suggest that elevated KCNIP3 levels in ALS patients may indicate stronger inhibitory control, suggesting a compensatory reduction in neuronal excitability.

Finally, ProtAIDe-Dx identified proteins predictive of PD that warrant further exploration. CSF NPTXR has emerged as a potential biomarker for synaptic dysfunction ^112,216,217^ and Lewy body disorders ^218,219^, and plasma levels were predictive of PD in our dataset. NPTXR regulates synaptic plasticity and excitatory synapse formation, and recent studies have shown it to have a neuroprotective role against PD pathology ^220,221^. SUMF1 has also been previously reported to be associated with PD^75^. The loss of SUMF1, which encodes an essential enzyme for activating cellular sulfatases ^222^, leads to lysosomal dysfunction and blocked autophagy ^223^. Therefore, SUMF1 may play a role in impaired α-synuclein clearance, given that lysosomal dysfunction contributes to the spread of α-synuclein ^224^. Finally, PRL demonstrates neuroprotective effects through activation of JAK/STAT and PI3K/Akt pathways and reduction of excitotoxicity in preclinical studies ^225^.

The architecture of deep learning models significantly influences their performance^226,227^. There are few systematic considerations needed when selecting architectures suited to specific biomedical tasks. Different data modalities often require distinct architectures. For example, convolutional neural networks (CNN) for imaging data and recurrent neural networks (RNN) for time-series data. Sample size is another key factor; transformer-based models have achieved remarkable success in recent years^228–232^, including in biomedical foundation models, but typically require very large samples sizes for effective training. Thus, selecting an appropriate architecture depends critically on both modality and sample size. In plasma proteomics, typically represented as unstructured tabular data, no consensus “gold standard” architecture has yet emerged. In this study, we adopted a MLP architecture, which achieved performance in neurodegenerative disease classification comparable to that reported in UK Biobank studies ^48,233^. We further benchmarked our MLP-based ProtAIDe-Dx against the state-of-the-art TabPFN model, pretrained on millions of synthetic datasets for general binary classification. ProtAIDe-Dx substantially outperformed TabPFN in predicting frontotemporal dementia (FTD) and achieved comparable performance across other disease categories. Given the extreme class imbalance for FTD (only 206 cases in GNPC), TabPFN may not have captured disease-specific signals, whereas our joint-learning strategy leveraged information across multiple neurodegenerative diseases to enhance FTD prediction. In summary, MLP-based models together with our joint-learning strategy, yielded slightly better performance under the current sample sizes. Nonetheless, considering the strong performance of TabPFN, joint-learning strategies, larger datasets, and pretraining on neurodegeneration-focused synthetic data may further improve transformer-based models for plasma proteomics.

The joint learning framework of ProtAIDe-Dx captured a mixture of disease-specific and more general pathological/neuroprotective signals. For instance, it extracted PD and ALS-discriminative signals using skeletomuscular enriched proteins, which reflect neuromuscular pathology ^234–236^. Another interesting finding is that ProtAIDe-Dx captured sex-related patterns shared with AD-discriminative signals, supporting the observation that biological sex may modulate vulnerability to neurodegenerative diseases ^237,238^. In addition to uni-disease signatures, ProtAIDe-Dx also captured some signals shared across multiple neurodegenerative diseases. For example, several embeddings were involved in neuronal cell death, inflammatory, or glial responses, as well as contributing to the discrimination of multiple neurodegenerative diseases. These signals are more likely to imply global neurodegenerative patterns rather than etiology-specific signatures ^239^. Interestingly, several embeddings were shared across PD and stroke/TIA. These signatures are involved in neuroinflammation ^240,241^ and vascular degeneration ^242,243^ but do not correlate with amyloid pathology. These signals also captured age and cognition effects. Whether these signals may reflect a co-pathology pattern ^244,245^ between PD and vascular diseases or simply a general ‘non-AD’ neurodegeneration axis distinct from AD pathology ^246^ remains to be explored. In addition to disease signatures, ProtAIDe-Dx extracted some potential brain health or resilience signals. These signals were enriched in multiple neuroprotective mechanisms, including neural integrity ^247^, immune/inflammatory response ^248,249^, metabolic maintenance ^250^, and synaptic plasticity ^251^. Consistent with these neuroprotective pathways, biomarker evidence also revealed that these signatures exhibit correlations with younger age, improved inflammation/atrophy markers, and better cognition. An interesting biomarker finding is that some signals were associated with more abnormal levels across almost all AD biomarkers, but were not associated with cognition, suggesting a potential role of resilience ^252^. Perhaps the most encouraging aspect of the proteomics embeddings is their generalizability and compactness. ProtAIDe-Dx compressed high-dimensional proteomics data into low-dimensional embeddings rich in neurodegeneration-related signals by classifying multiple neurodegenerative diseases. This suggests that the embeddings could be extended to new neurodegeneration-related tasks ^253,254^ and potentially to previously under-studied neurodegenerative diseases ^255^.

In all, ProtAIDe-Dx showed great promise in disease diagnosis, probing disease heterogeneity, identifying novel proteins of interest, and generating health- and disease-related signatures. However, there are still challenges that we wish to highlight to facilitate future studies exceeding the performance of ProtAIDe-Dx in real-world samples. A major challenge of ProtAIDe-Dx is that its diagnostic accuracy has not yet reached the level required for standalone clinical use. This limitation may be due to several factors. The performance of ProtAIDe-Dx was comparable to that of other studies using UK Biobank data ^47,48,163^, suggesting that there may be a performance ceiling of plasma proteomics as biomarkers. Another factor might be a limited set of specific aptamers that target arbitrary protein conformations, which are also secreted or surface-based and are detectable in blood. This is a particular challenge for brain diseases, since many disease-relevant proteins are likely brain-expressed and many of these do not cross the blood-brain barrier. Additionally, plasma p-tau biomarkers have achieved much better performance discriminating AD than we found here ^7,256^ due to the identification of specific peptides and post-translational modifications directly related to disease pathology. To achieve similar improvements, mass spectrometry and/or other approaches may be needed for more comprehensive screening of peptides and protein fragments ^257^. Another challenge of ProtAIDe-Dx is its relatively poor site generalization performance, which hinders its applicability to new sites. This limitation may be due to several factors. Strong site effects in plasma proteomics ^258^ cannot be ignored and may be mitigated by statistical harmonization techniques ^169,170,259^. In addition, demographic, cognitive, disease distributions, or even diagnoses by clinicians varied across cohorts, potentially introducing distribution bias that hampers generalization. This issue could be addressed using adversarial approaches to unlearn distributional bias ^260,261^. As discussed, the training labels may not be sufficiently reliable due to the lack of biomarker support and imperfect harmonization of diagnostic criteria across cohorts. To address this issue, integrating plasma proteomics studies conducted during life with neuropathological assessments at death may provide a practical solution for enabling model training with ground truth diagnostic labels. Alternatively, future studies are recommended to incorporate more biomarker-confirmed samples to enable models to more accurately capture biologically meaningful signals for disease classification. Given the remarkable success of unsupervised learning approaches (e.g., disease subtyping^200,262^) in neurodegenerative research, another promising pursuit would be to apply unsupervised learning on proteomics data, as disease-relevant variability within the population may only partially overlap with clinical diagnoses yet could provide valuable insights into the underlying biology. Another challenge is presented by the potential confounding effects of various factors on protein levels. For example, medication use can significantly affect circulating protein levels, sometimes exceeding normal physiological ranges ^74^, and may therefore dominate model predictions. Our finding of ACHE predicting AD is likely driven by acetylcholinesterase inhibitors, a common treatment for MCI and AD, elevating plasma ACHE levels specifically in these patients, and the story is similar with KCNIP3 levels in ALS patients on riluzole. Another major confounding factor is aging, which is closely linked to the onset of neurodegenerative diseases and which strongly affects proteomic expression levels ^263^. These concerns could be mitigated by incorporating covariates or using conditional prediction strategies in the model design, which is worthwhile to explore in future work. Hopefully, many of these challenges can be addressed or improved in future releases of GNPC^124^.

In summary, ProtAIDe-Dx represents a pioneering attempt toward the development of scalable, minimally invasive, and multi-disease diagnostic tools for neurodegenerative diseases. Leveraging deep neural networks trained on the largest neurodegeneration-focused plasma proteomics dataset to date (N = 17,187; 7,595 proteins), the model achieved modest out-of-site performance across six different conditions. Beyond classification accuracy, ProtAIDe-Dx revealed shared molecular signatures across clinical diagnoses, identified a compact set of proteins predictive of multiple diseases, produced low-dimensional proteomics capturing disease- and resilience-related signals, and generated diagnostic probabilities that were correlated with PD symptom progression, CSF biomarkers, and imaging data in a separate memory clinic sample. Furthermore, the learned proteomic embeddings improved diagnostic accuracy beyond what was achievable using accessible clinical biomarkers alone. Despite its promise, predictive proteomics as a field faces several challenges, including insufficient diagnostic accuracy for clinical deployment, limited generalizability across sites, and potential confounding effects. Future work should address these limitations by incorporating more reliable diagnostic labels to improve accuracy, refining model architecture to enhance generalization, and implementing strategies to mitigate confounding biases. Overall, ProtAIDe-Dx establishes a robust benchmark for AI-driven proteomics tools, paving the way for precision medicine in neurodegenerative diseases.

## Supporting information

SuppData

## Acknowledgements

This work was supported by the SciLifeLab & Wallenberg Data Driven Life Science Program (grant: KAW 2020.0239), the Crafoord Foundation (20230790), the Swedish Research Council (2024-03642) and the US National Institute of Health (U01 AG079847-02). The BioFINDER study was supported by the National Institute of Aging (R01AG083740), European Research Council (ADG-101096455), Alzheimer’s Association (ZEN24-1069572, SG-23-1061717), GHR Foundation, Swedish Research Council (2022-00775, 2021-02219), ERA PerMed (ERAPERMED2021-184), Knut and Alice Wallenberg foundation (2022-0231), Strategic Research Area MultiPark (Multidisciplinary Research in Parkinson’s disease) at Lund University, Swedish Alzheimer Foundation (AF-980907, AF-994229), Swedish Brain Foundation (FO2021-0293, FO2023-0163), WASP and DDLS Joint call for research projects (WASP/DDLS22-066), Parkinson foundation of Sweden (1412/22), Cure Alzheimer’s fund, Rönström Family Foundation, Konung Gustaf V:s och Drottning Victorias Frimurarestiftelse, Skåne University Hospital Foundation (2020-O000028), Regionalt Forskningsstöd (2022-1259) and Swedish federal government under the ALF agreement (2022-Projekt0080, 2022-Projekt0107). The precursor of 18F-flutemetamol was sponsored by GE Healthcare. The precursor of 18F-RO948 was provided by Roche. Our computational work was supported by workspace mms-proteomics-kb from the Alzheimer’s Disease Data Initiative (https://www.alzheimersdata.org), project sens2023026 by the National Academic Infrastructure for Supercomputing in Sweden (NAISS; https://www.naiss.se) at UPPMAX, project NAISS 2024/22-457 by Chalmers e-Commons at Chalmers, and the Berzelius resource (Berzelius-2025-231) funded by the Knut and Alice Wallenberg Foundation at the National Supercomputer Centre. NAISS is partially funded by the Swedish Research Council through grant agreement no. 2022-06725. The funding sources had no role in the design and conduct of the study; in the collection, analysis, interpretation of the data; or in the preparation, review, or approval of the manuscript.

## Method

### Datasets

#### Global Neurodegenerative Proteomics Cohort (GNPC)

Global Neurodegenerative Proteomics Cohort (GNPC, https://www.neuroproteome.org/), launched in 2023, is a mega consortium combining multiple dementia and population cohorts, including health aging, Alzheimer’s Disease (AD), Parkinson’s Disease (PD), frontotemporal dementia (FTD), amyotrophic lateral sclerosis (ALS), and stroke/TIA ^124^. The latest GNPC v1.3MS release collected 20,532 participants from 22 contributors (sites), 3,950 of whom had longitudinal visits. We used baseline visit proteomics data for model development.

In this study, we selected 17,187 participants based on SomaLogic 7k proteomics availability. Site U’s proteomics data were from serum, but we kept site U to maximize the sample size and learn modality-agnostic signatures. For the 9,708 participants that were neither diagnosed as Control, nor with any of the five above mentioned diseases, we visualized the distribution of these participants (Fig. S18). Based on MMSE and CDR, we mapped participants with good cognition (MMSE >= 26 or CDR=0) as “Control”, participants with mild cognitive impairment (20<=MMSE <= 25 or CDR=0.5) as “MCI-SCI”. Among the remaining 1,606 participants, 1,062 participants with poor cognition (MMSE <= 10 or CDR>=1) were labeled as “ComputedDementia”, and 542 participants without valid MMSE or CDR were labeled as “Unknown”. Table S1 shows the demographics and cognition (Mini-Mental State Examination, MMSE) distribution of 17,187 participants by site, and the corresponding distribution of clinical diagnoses can be found in Table S2. Race and ethnicity information can be found in Fig. S19. It is noted that diagnoses of some sites were confirmed by biomarkers, but not for most sites. In the following analysis, the patients with either Stroke or TIA were labeled with Stroke in figures for ease of visualizations, but are referred to stroke/TIA in the Main Text.

All GNPC blood samples were shipped separately by each contributor and were analyzed by SomaLogic, coordinated by Gates Ventures. SomaLogic applied slow off-rate modified aptamer (SOMAmer) technology, in which chemically modified nucleotides enable high-specificity and high-affinity binding to target proteins. The resulting proteomic data were standardized, normalized, and calibrated, with protein abundances reported in relative fluorescent units (RFU). Before integration into the GNPC cohort dataset, aptamers were mapped to UniProt ^124^. We removed the outlier values for each protein that exceeded six standard deviations.

#### BioFINDER-2 cohort

The Swedish BioFINDER-2 dataset (https://biofinder.se/two/, NCT03174938) is a prospective cohort in the south of Sweden spanning the full continuum of AD as well as including patients with non-AD neurodegenerative diseases. Participants are deeply phenotyped, including clinical assessment, CSF/blood sampling, PET, and MRI imaging data. BioFINDER-2 is a participating site in GNPC, but in a sub-analysis aiming to evaluate the disease probability predicted by the different models in relation to key markers of AD and neurodegenerative diseases, we focused specifically on this cohort. For the BioFINDER-2 sub-analysis, we selected 1,786 participants with plasma SomaLogic 7k proteomics data. The demographics and cognition distribution of 1,786 participants can be found in Table S12.

We grouped participants into 6 different groups: Cognitively unimpaired (CU), SCD, MCI, AD, Parkinsonism and Other diseases. The BioFINDER-2 dataset inclusion and exclusion criteria have been described in detail previously ^264^. Briefly, CU participants needed to have a MMSE of at least 27 (if < 66 years old) or 26 (if >= 66 years old) and no signs of cognitive symptoms as assessed by physicians specialized in cognitive disorders. Participants with SCD or MCI were all referred to a memory clinic due to cognitive symptoms, had an MMSE between 24 and 30 and did not fulfill criteria for any dementia according to the DSM-5. Participants were classified as MCI if they performed at least 1.5 standard deviations below the normative score on at least one cognitive domain from an extensive neuropsychological test battery ^12^, while SCD participants performed better than - 1.5 standard deviations. Patients with dementia fulfilled the DSM-5 criteria for dementia and all patients with AD dementia were Aβ-positive (based on CSF Aβ42/Aβ40). Patients with non-AD neurodegenerative diseases were also included in the cohort. Patients with PD, dementia with Lewy bodies, atypical parkinsonism disorders formed the group “Parkinsonism”. Patients with other diseases were grouped together (labeled “Other”) and included patients with FTD spectrum disorders, vascular dementia, and one patient with etiology not otherwise specified. Clinical diagnosis of AD dementia or other neurodegenerative diseases was determined by experienced clinicians.

To benchmark generalization performances on BioFINDER-2 (Fig. 4B) against leave-one-site-out on GNPC, we performed additional diagnostic grouping to match GNPC based on clinical diagnosis and biomarkers of BioFINDER-2. Participants with clinical diagnosis as Normal were labeled with Control; patients with abnormal CSF Aβ42/Aβ40 ratio and clinical diagnosis as AD are labeled as AD; patients with a clinical diagnosis with PD, DLB, or Parkinsonism (not otherwise specified) were labeled with Parkinsonism Disorder (PD); patients with a clinical diagnosis behavioral variant FTD, semantic variant of primary progressive aphasia, FTD (not otherwise specified) or a SCD with a MAPT mutation were labeled with FTD; patients with infarcts were labeled with Stroke. It is noted that stroke is not exclusive to other diagnosis categories. For example, an AD patient might also be diagnosed with Stroke. In summary, we got 609 control participants, 261 AD patients, 135 PD patients, 43 FTD patients and 117 stroke patients.

#### Biomarkers of interest in BioFINDER-2

Multiple biomarkers and the MMSE as a global measure of cognition were investigated in BioFINDER-2. The AD biomarkers were Aβ status (based on CSF Aβ42/Aβ40) and tau-PET standardized uptake value ratio (SUVR) in a temporal meta-ROI (tracer ^18^F-RO948) ^265,266^. The positivity of CSF p-tau217 was determined at a cutoff of 11.42 pg/ml ^12^. Structural MRI markers of interest were the cortical thickness in an AD signature comprised of temporal lobe regions ^155^, whole-brain cortical thickness, and ventricular volume (average of lateral ventricle volume in both hemispheres divided by total intracranial volume). T1-weighted MRI were processed with FreeSurfer v6.0. White matter hyperintensity burden (divided by total intracranial volume) was measured based on FLAIR and T1 images processed with Sequence Adaptive Multimodal SEGmentation (SAMSEG) tool from FreeSurfer v7.1. α-synuclein status was available from seed amplification assay done in CSF, as described previously ^12^. For a subset of participants in the Parkinsonism group (n=100), the Unified Parkinson’s Disease Rating Scale (UPDRS) was also available.

#### Model cross-validation and generalization

After diagnostic label mapping, 11,803 participants were labeled as either Control or one of the five aforementioned diseases, 662 participants diagnosed with AD but exhibiting healthy cognitive scores (MMSE >= 26) were labeled as “HealthyAD”, 3,116 participants were labeled as “MCI-SCI”, 1,064 as “ComputedDementia” with poor cognitions (MMSE < 19 or CDR>=1), and 542 as “Unknown”. The 11,803 participants in the Control and disease groups were used as the development set for model training and evaluation, including 10-fold cross-validation and leave-one-site-out procedures. The “HealthyAD”, “MCI-SCI”, “ComputedDementia”, and Unknown” groups were held out as additional test sets. We also conducted a supplementary experiment by including MCI-SCI patients in model development (Fig. S7).

We conducted a 10-fold cross-validation procedure to evaluate the performances of ProtAIDe-Dx models on GNPC. Participants from each site were evenly split into 10 folds, so each fold contained data from all sites. A 9-1-1 train-validation-test split was employed, with 8 of 10 folds as a training split, one fold as a validation split, and the remaining one as a test split. This process was repeated 10 times, with each fold serving as the test split once.

To test the site generalization performance of the proposed ProtAIDe-Dx models, a leave-one-contributor-out scheme was employed. Data from one site was reserved as the test split, while the data from the remaining contributors served as a training-validation split.

The sites used as test sets were selected based on the following criteria: for any of the six conditions, 1) this site had 200 or more participants with non-missing diagnoses; 2) this site had at least 5 participants with minor diagnosis categories for this condition. In total, 14 testing sites fit this criteria. Detailed information can be found in Table S13. Notably, the BioFINDER-2 cohort was part of GNPC, therefore, we excluded BioFINDER-2 participants when training and tuning the ProtAIDe-Dx model for testing on BioFINDER-2.

### Machine learning model development

#### Feature selection

A feature selection procedure was employed to reduce the number of input proteins. On each training and validation split, we conducted both GLM association analysis and XGBoost ^52^ predictive analysis to select informative proteins as input features. Since GLM does not require any hyperparameters to set, we merged training and validation participants together to run the GLM association analysis for each of the six conditions and each of the 7,595 SomaLogic 7k proteins following:

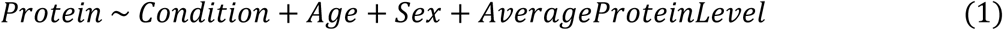

*AverageProteinLevel* is the average protein level across all 7k proteins to control for individual expression differences ^267^.

After running 7,595 GLMs for each target, we first selected proteins with fold change of beta *Condition* larger > 2 or smaller than 0.5 and then picked the top 5 proteins with the smallest corrected p values of beta *Condition*.

We also ran XGBoost predictive models on each training and validation split to select predictive proteins. We merged training and validation participants together and re-split these participants into 10 subfolds, 9 used for training models and 1 as validation split to tune model hyperparameters. This procedure was repeated 10 times to predict each condition, with each subfold serving as the validation split once. After all 10 models for each condition were trained, proteins internally selected for building models by all 10 XGBoost models were kept.

In this way, the number of input proteins was reduced from 7,595 to 200 - 300 proteins as input features, which vary across each train-val-test split. In total, 648 proteins were kept across 10 train-validation-test splits. This feature selection procedure was performed for both cross-validation and leave-one-contributor-out experiments.

#### Classification metrics

Accuracy is an inappropriate and potentially misleading metric given the imbalanced distribution across the six conditions, we therefore chose balanced classification accuracy (BCA) as classification metric, which was commonly used in AD classification tasks ^268,269^. We also reported Area Under Curve (AUC) scores for reference.

#### Random Forest as baseline approach

Random Forest models were employed as baseline machine approaches to classify binary targets separately. We therefore trained six Random Forest models for classifying six binary conditions. The input proteins to Random Forest models were z-normalized by mean and standard deviation computed from the training split. To get optimal validation prediction accuracies, a grid search on hyperparameters, including maximum tree depth, number of features for best split, and probability threshold, was performed on the validation split. The Random Forest model trained with optimal hyperparameters was then applied to the test split. This procedure was repeated for each training-validation-test split, including cross-validation and leave-one-site-out.

#### XGBoost as baseline approach

XGBoost models were employed as baseline machine approaches to classify binary targets separately. We therefore trained six XGBoost models for classifying six binary conditions. The input proteins to XGBoost models were z-normalized by mean and standard deviation computed from the training split. To get optimal validation prediction accuracies, a grid search on hyperparameters, including maximum tree depth, subsampling of training data, and probability threshold, was performed on the validation split. The XGBoost model trained with optimal hyperparameters was then applied to the test split. This procedure was repeated for each training-validation-test split, including cross-validation and leave-one-site-out.

#### TabPFN as baseline approach

TabPFN models were employed as baseline machine approaches to classify binary targets separately. We therefore fitted six TabPFN models based on pretrained TabPFN classifier for classifying six binary conditions. We performed several additional postprocessing steps before proteomics data was input into deep learning models, which were same for both TabPFN and ProtAIDe-Dx. First, each participant’s proteomics values were normalized by their averaged protein levels ^267^. Second, we fitted a 10-nearest neighbor data imputer onto the training split to impute missing protein entries. Third, a Gaussian rank normalizer ^270^ was fitted on training split to ensure normalized proteomics are following a normal distribution. The optimal validation probability threshold was selected based on best F1 scores on validation set. The TabPFN model fitted with optimal probability threshold was then applied to the test split. This procedure was repeated for each training-validation-test split, including cross-validation and leave-one-site-out.

#### ProtAIDe-Dx

The ProtAIDe-Dx models were implemented as multiple layer perceptron (MLP)-based networks. Input proteins were fed into multiple multilayer neural networks to classify six binary conditions jointly. The key consideration of choosing multi-task over multi-class approach for ProtAIDe-Dx is that diagnostic labels are often incomplete in GNPC, as the dataset was aggregated from multiple cohorts with varying research objectives. For example, a participant diagnosed with AD may not have been formally assessed for PD, resulting in missing labels for certain conditions. In a multi-class classification framework using one-hot label vectors across six diagnostic categories, many vectors would contain missing values, necessitating either imputation or exclusion of these samples, both of which would usually be suboptimal. By contrast, a multi-task learning framework maximizes sample utilization, as each task is trained independently on subjects with labels available for that specific condition.

The imbalanced distribution of the six conditions may bias the model toward the majority class and lead to poor generalization if the loss function is not carefully designed. The loss function of the proposed ProtAIDe-Dx model is a weighted combination of binary cross-entropy loss *L_BCE_* and multi-class rank loss ^271^ *L_RL_*. Label smoothing mitigates overfitting to potentially noisy clinical annotations by calibrating model confidence, while rank loss enhances robustness to distributional skew by optimizing relative sample rankings rather than absolute probabilities. This combined approach ensures improved generalization across all conditions despite uneven representation.

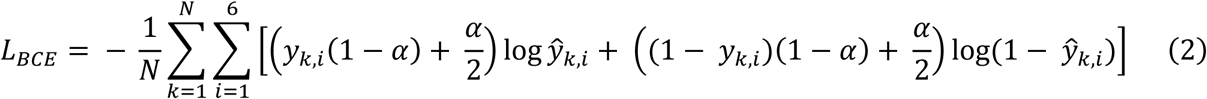

Where *N* is the number of participants, *k* is the participant index, *i* is the index of conditions, *y_k,i_* is the true label for the participant *t* and condition *i*, *a* is a hyperparameter label smoothing factor to set, 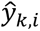 is the model predicted probability for the participant *k* and condition *i*.

The role of rank loss *L_RL_* is to constrain the rank of predicted probabilities across conditions, enabling better jointly learning of information across targets. For any two conditions *i* and *j*, the rank loss *L_RLi_^,j^* is

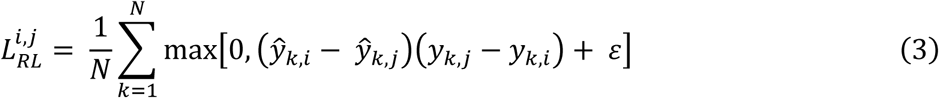

Where *N* is the number of participants, *k* is the participant index, *i* and *j* are indexes of two conditions. 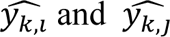 are predicted participant *k*’s probabilities for condition *i* and *j*. ε is a hyperparameter set as 0.25.

The rank loss across all conditions is

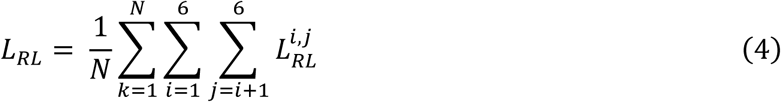

Therefore, the overall loss function for the proposed ProtAIDe-Dx model is

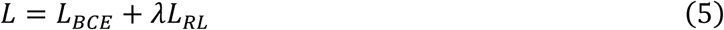

Where λ is a hyperparameter to control the weight of *L_RL_*

The probability threshold for each target was determined for the highest validation F1 score. We performed a hyperparameter search on the validation split to get the optimal validation balanced accuracy (BCA) using Optuna ^272^ with 50 trials. The optimal validation probability threshold was selected based on best F1 scores on the validation set. The search range of hyperparameters can be found in the following Table S14, with optimal searched hyperparameters listed in Table S15 (model evaluated on BioFINDER-2) and SuppData 10 (all cross-validation and leave-one-site-out models). An illustration of ProtAIDe-Dx architecture can be found in Fig. S20.

We performed several additional postprocessing steps before proteomics data was input into ProtAIDe-Dx models. First, each participant’s proteomics values were normalized by their averaged protein levels ^267^. Second, we fitted a 10-nearest neighbor data imputer onto the training split to impute missing protein entries. Third, a Gaussian rank normalizer ^270^ was fitted on training split to ensure normalized proteomics are following a normal distribution.

#### Proposed Ensemble of XGBoost and ProtAIDe-Dx

We conducted an ensemble approach for the developed XGBoost and ProtAIDe-Dx models. The ensemble approach’s output probability for each condition was a weighted sum between XGBoost and ProtAIDe-Dx: 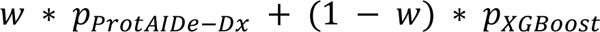. On the validation split, we searched for the optimal weight *w*, ranging from 0 to 1 with the step as 0.01, to get highest validation balanced accuracy. This procedure was repeated for each training-validation-test split, including cross-validation and leave-one-site-out.

#### Model generalization to new sites by k-shot transfer learning

We developed a k-shot transfer learning framework ^173,273,274^ to help the developed model generalize to new tasks or new sites. Taking generalization to new sites as an example, only K participants were needed to transfer the ProtAIDe-Dx model to this new site, which makes ProtAIDe-Dx easy to scale to real-world clinical settings given model calibration^275,276^ only needs K participants. In this study, we picked K=100 to balance available sample size for each condition and generalization performance. First, the ProtAIDe-Dx model was applied to new sites to get proteomic embeddings. Second, a simple logistic regression model was trained on K participants’ proteomics embedding and tested on the remaining participants. The performances were evaluated on remaining participants. Similarly, the developed ProtAIDe-Dx model could transfer to new neurodegeneration-related tasks by training a simple machine learning model on K participants.

The underlying assumption is that ProtAIDe-Dx’s embeddings have captured enough signals to represent neurodegenerative diseases broadly. Therefore, training a simple machine learning model on low-dimensional proteomics embedding with limited subjects will avoid overfitting issues compared to directly training new models on raw proteins.

#### Estimating predictive importance

To estimate the predictive importance of individual proteins, the PermFIT approach ^59^ was adopted on each test split. For each input protein, the PermFIT approach randomly permuted this protein across participants and computed the cross-entropy differences compared to unpermutated data. The mean and standard deviation of cross-entropy differences for this protein were obtained from 100 permutations, and p-values were calculated based on the mean and standard deviation, assuming a normal distribution.

In this study, we measured the predictive importance of proteins in two ways, including Fold Count and Z statistics. Once PermFIT was done on all 10 splits, the Fold Count approach counted the frequency of important proteins across those splits using FDR-corrected p values. The Z statistics approach computed the z scores (mean divided by standard deviations) on each test split and then averaged z scores across 10 splits to get the predictive importance of proteins.

To estimate the importance of individual model embeddings in predicting specific diagnoses, we used the Haufe approach. Haufe and colleagues ^277^ proved that covariance is more reliable than weights (betas) to interpret feature importances in linear models. Given the embeddings were linearly combined to make predictions, we computed the covariance between embeddings and predicted probabilities for each condition to estimate the embeddings’ feature importance.

#### Model evaluation on GNPC

In addition to reporting classification accuracy, we performed multiple experiments to validate ProtAIDe-Dx models on GNPC data. First, we correlated predicted AD probability with MMSE scores. Given that each train-validation-test split had different probability thresholds, we first divided AD probability by corresponding AD probability threshold for normalization. Then, we took the logarithm of the mean AD probabilities of participants with the same MMSE score on each testing split. Finally, we computed the Pearson correlation between MMSE scores and the logarithms of the mean AD probabilities. Similarly, we computed the logarithms of mean AD probabilities by *APOE* ε2/ε4 carrier group on each test split, then averaged them across 10 splits.

#### Model generalization to new task: prediction of longitudinal clinical progression

We validated the generalization ability of nonlinear proteomics embeddings to new tasks. More specifically, we took the embeddings from baseline visits and used these to predict whether participants with CDR 0 at baseline visits would progress to CDR 0.5 or 1 in their following visits or not. For 3,942 participants with longitudinal visits, we selected participants with CDR 0 at baseline visits and non-decreasing CDR during the following visits. For example, participants with CDR 0, 1, 0 in their first, second, and third visits would be excluded. In total, we selected 1,945 participants with stable CDR 0 in their following visits and 107 participants with progressive CDR in their following visits. It is noted that the baseline visits of the 1,945+107 = 2,052 participants have been used in the 10-fold cross-validation procedure for ProtAIDe-Dx development. We trained Logistic Regression (LR) models to predict whether these 2,052 participants would progress to CDR 0.5 or 1 in their following visits or not, following the former 10-fold cross-validation procedure. We adopted default hyper-parameters for logistic regression models from the sklearn package ^278^.

#### Dimensionality reduction of predicted probabilities with tSNE

We selected 6,124 participants who were either diagnosed as Recruited Control (N=1,095), AD (N=1,889), PD (N=2,284), FTD (N=176), ALS (N=435), or stroke/TIA (N=335) to project onto a 2-dimensional probability map. We excluded subjects with all negatives or multiple positives for the six targets. It is noted that the predicted probabilities of the 6,124 participants were all from test splits, following the former 10-fold cross-validation procedure.

The dimensionality reduction algorithm was chosen as tSNE from the OpenTSNE library ^279^. We used OpenTSNE over sklearn because OpenTSNE supports inference on new data with fitted tSNE models, allowing us to investigate new participants such as MCI-SCI patients on the same tSNE map. The parameters of tSNE were default values except for setting perplexity to 1000 to better capture global structures.

#### Assessing disease heterogeneity

For each target, KMeans clustering was performed on the 2-dimensional tSNE probability maps of participants within each diagnostic category. To pick the optimal number of clusters, we looped it over from 2 to 10 to pick the one with the highest Silhouette score. In summary, we got 2 Control clusters, 3 AD clusters, 4 PD clusters, 2 FTD clusters, 2 ALS clusters, and 5 stroke/TIA clusters.

To figure out the differentially expressed proteins across clusters, we ran GLMs across 7k proteins on positive participants for each diagnostic category. Therefore, for each diagnostic category, we have GLMs following the formula:

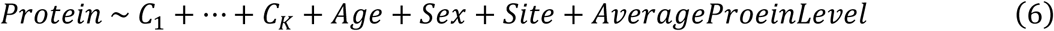

*C_1_*… *C_k_* are the binary variables indicating which cluster the participants were from. *Site* is the categorical variable that indicates which site the participants were from.

#### Assessing site-to-site generalization variability

We conducted a site-to-site generalization experiment in which models were trained exclusively on data from a single site (development site) and evaluated on all remaining sites (test sites). As expected, classification performance was higher on the development site than on the test sites. We selected TabPFN as the predictive model because it does not require hyperparameter tuning. The same sites used in the leave-one-site-out experiments were included here. For each development site, the data were randomly split into two equal halves. Two TabPFN models were trained independently on each half and evaluated on the other half, with development-site accuracy defined as the mean of these two within-site accuracies. The two trained TabPFN models were then applied to the remaining external test sites, and test-site accuracy was defined as the mean performance of both models on the test sites. To quantify site-to-site variability, we defined relative performance as the ratio of test-site accuracy to development-site accuracy, with lower values indicating stronger site effects.

#### Enrichment analysis

To evaluate the cell-type expression of genes of interest, we utilized two independent single-cell RNA sequencing (scRNA-seq) datasets. The first dataset comprised single-cell transcriptomes from 2.3 million cells obtained from the aged human prefrontal cortex of 427 participants in the Religious Order Study (ROS) and the Rush Memory and Aging Project (MAP) ^280^. This dataset includes neuronal, glial, and vascular cell types. The second dataset, the Human Brain Vascular Atlas, profiled mainly vascular and perivascular cell types, using 143,793 single-cell transcriptomes from the hippocampus and cortex of eight post-mortem samples ^281^.

For the two datasets, we downloaded the Seurat objects and used the R package Seurat v4.3.0 ^282,283^, applying the AverageExpression function to compute cell-type expression levels. We then calculated the proportion of gene expression across all major neuronal and non-neuronal populations.

For organ enrichment analysis, we used organ-enriched genes as defined in previous studies ^284^ based on the Gene Tissue Expression Atlas (GTEx) bulk RNA-seq database. A gene was classified as organ-enriched if its expression was at least four-fold higher in a specific organ or tissue compared to any other, following the Human Protein Atlas definition ^285^. We considered organ categories including adipose tissue, artery, brain, esophagus, heart, immune, intestine, kidney, liver, lung, muscle, pancreas, skin, stomach, and whole blood. Cell- and organ-enriched genes were then mapped to proteins quantified in the SomaScan assay.

To determine whether a specific set of proteins exhibited preferential expression in certain cell types, we calculated the average expression level of the significant protein list across each cell type. To determine whether this enrichment exceeded random expectation, we generated 10,000 randomly sampled gene lists from the Somalogic background set, each maintaining the same protein count as the significant list. A probability distribution was then constructed based on the average expression levels of these random lists across cell types. This allowed us to quantify the likelihood that the observed enrichment was greater than expected by chance, while accounting for background expression variability. Cell types with a false discovery rate (pFDR)<0.05 (Benjamini–Hochberg correction) were considered significantly enriched.

We performed overrepresentation analyses of KEGG pathways and Gene Ontology (GO) Biological Process (BP) terms using enrichKEGG() and enrichGO() functions in clusterProfiler (v4.14.4)^286^ and org.Hs.eg.db (v3.20.0)^287^ in R (v4.4.2). The input gene lists consisted of proteins that were significantly up-, down-regulated or both in disease clusters defined by logFoldChange and false discovery rate (pFDR)<0.05 (Benjamini–Hochberg correction). We used all human proteins as the background set for overrepresentation analysis. We reported only those KEGG pathways and GO BP terms that met a false discovery rate (FDR) threshold of <0.05. To reduce redundancy among significantly enriched BP terms, we applied the clusterProfiler simplify() function using Wang’s semantic similarity measure with a cutoff of 0.7.

Enrichment analyses were performed on proteins differentially expressed across tSNE clusters. Enrichment analysis was additionally performed on embedding-specific proteins. To annotate the biological process of each nonlinear proteomics embedding, we found “embedding specific” proteins using regression. For each embedding, we took the 648 proteins from the cross-validation feature selection to train an XGBoost regression model, with grid search on validation split for the lowest mean squared errors. It is noted that the model was trained to generalize to BioFINDER-2 has 32 embedding dimensions. Once these models were trained, we thresholded proteins with ‘total_gain’ over 50 as “embedding specific” proteins for enrichment analysis.

#### Model evaluation on BioFINDER-2

In the BioFINDER-2 dataset, where we correlated each biomarker with the different predicted probabilities separately in each of the six diagnostic groups (CU, SCD, MCI, AD, Parkinsonism, Other). FDR correction was applied across all comparisons performed. We also calculated the Pearson correlation between embeddings and biomarkers for BioFINDER-2 participants who had corresponding biomarker available. FDR correction was applied across all comparisons performed.

#### Differential diagnostics of multiple neurodegenerative diseases

We further investigated whether proteomics embeddings would provide unique or additive value in distinguishing multiple neurodegenerative diseases in the BioFINDER-2 dataset. We selected 231 AD patients (with abnormal CSF Aβ42/Aβ40 ratio), 111 PD patients (with positive CSF α-synuclein status), 39 FTD patients, and 20 Stroke patients (with infarcts), excluding patients with multiple neurodegenerative diseases. We performed a five-fold cross-validation, ensuring the diagnosis distribution was balanced across folds. Three out of five folds were used to train an SVM model, one fold was used to tune hyperparameter C from 0.001 to 100 to get an optimal balanced accuracy score, and the remaining one was used for the test. This procedure was repeated five times to ensure each fold was used as a test fold.

Four sets of features were selected to validate the effectiveness of our proteomics embeddings. Model 0 only used age and sex; Model 1 used age, sex, and the top 5 principal components of proteomic embeddings; Model 2 used age and sex plus clinical biomarkers, including MMSE, plasma p-tau217, plasma NEFL, and mean cortical thickness of AD-signature meta-ROI; and Model 3 used all features in Model 0, Model 1 and Model 2. It is noted that age, MMSE, mean cortical thickness of AD-signature meta-ROI, plasma p-tau217 and plasma NEFL were z-normalized. Both PCA and z normalization were fitted on training splits. Once models were trained, we concatenated participants from five test folds to perform bootstraps 1000 times to perform statistical tests.

#### Mixed-effect modeling of longitudinal cognitive decline

To assess whether baseline characteristics (e.g., clinical diagnoses, model predictions) could differentiate longitudinal trajectories of decline, we fitted linear mixed-effects models with individual variability modeled as random effects. Cognitive performance was measured using the MMSE.

First, we evaluated whether baseline clinical diagnoses could differentiate longitudinal trajectories of decline.

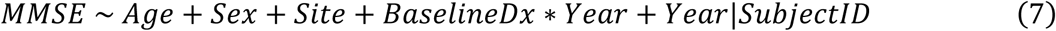

*BaselineDx* is the categorical variable that indicates baseline diagnoses, *Year*|*SubjectID* specifies random effects of individuals over time (in units of years).

Then we evaluated whether baseline predictions could differentiate longitudinal trajectories of decline, we included baseline diagnoses as covariates.

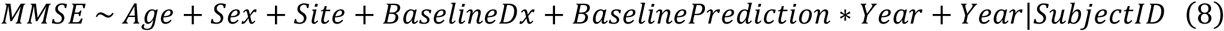

*BaselinePredition* is the categorical variable that indicates baseline predictions, *Year*|*SubjectID* specifies random effects of individuals over time (in units of years).

#### A two cut-off approach of predicted probabilities

In the external BioFINDER-2 dataset, we applied a two cut-off strategy on predicted probabilities to determine biomarker positivity. Participants were divided into non-SCD and SCD groups, with the non-SCD group used to derive the cut-offs and the SCD group serving as the test set. Cut-offs were chosen to yield 90% NPVs and PPVs, except for CSF α-synuclein, where the PPV was set at 40% due to sample validity constraints.

#### Individual disease risk report

To assess contributing proteins for individual prediction, SHapley Additive exPlanations (SHAP) values ^288^ were computed for each of six conditions, with training participants as the background set. To associate the top contributing proteins with health-related traits, we applied a proteome-phenome atlas^49^ to find the top correlated traits for each protein. The proteome-phenome atlas was computed based on Olink proteomic data. Therefore, there might be no matched results for some of the contributing proteins; we only included available protein-trait associations.

#### Statistical analysis

We applied a corrected resample t-test to compare model performances in cross-validation and leave-one-site-out experiments. To examine whether the distribution of variables varies across tSNE clusters or not, we conducted two-proportion z tests for binary variables with two clusters, chi-square tests of independence for both binary variables with three or more groups and multi-category variables, t-test for continuous variables with two clusters, and one-way ANOVA for continuous variables with three or more groups. To compare model performances in comorbidity detection, we applied t-tests on 1000 bootstrapped accuracies. We conducted t-tests for comparing probability distributions across biomarker status. It is noted that all p values were FDR corrected with *a* as 0.05 using Benjamini–Hochberg procedure.

## Data Availability

GNPC (https://www.neuroproteome.org/) will be open-acess upon the publication of this study. Pseudonymized BioFINDER-2 data will be shared by request from a qualified academic investigator for the sole purpose of replicating procedures and results presented in the article and as long as data transfer is in agreement with EU legislation on the general data protection regulation and decisions by the Swedish Ethical Review Authority and Region Skåne, which should be regulated in a material transfer agreement.

## Code Availability

Code for the ProtAIDe-Dx model development/evaluation/biological annotations can be found at https://github.com/DeMONLab-BioFINDER/An_ProtAIDe-Dx. Co-author (YX) reviewed the code before merging it into the GitHub repository to reduce the chance of coding errors.

