## Supplementary for "Benchmarking the AI-based diagnostic potential of plasma proteomics for neurodegenerative disease in 17,187 people"

### Supplementary Figures

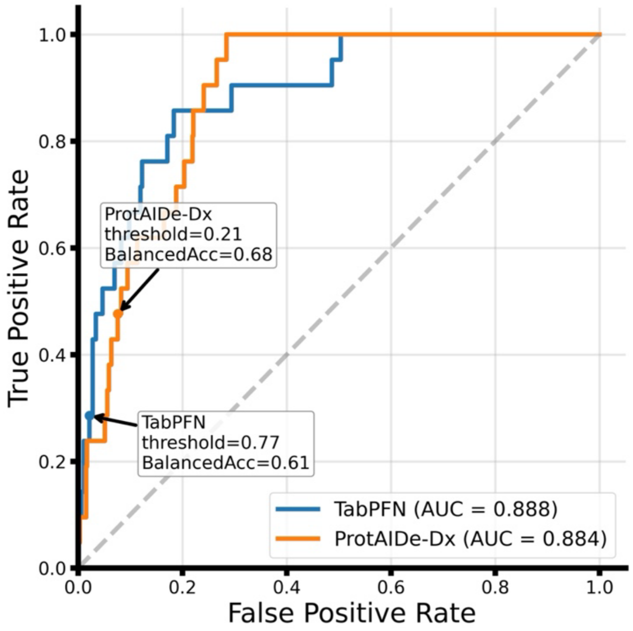


**Figure S1. AUROC can be misleading under extreme class imbalance.** ROC curves for ProtAIDe-Dx and TabPFN in one test fold's FTD prediction (1% positive) show similar AUROC, whereas ProtAIDe-Dx yields substantially higher balanced accuracy. Decision thresholds were selected by maximizing F1 on the validation set using the same procedure for both models.

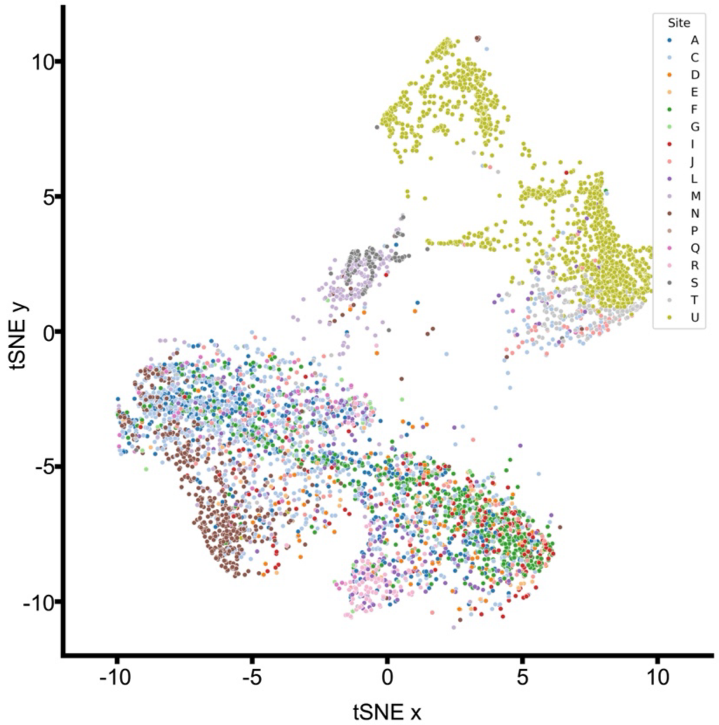


**Figure S2. Site distribution of 6,332 test participants on tSNE diagnostic probability map.** The test participants and tSNE were the same as in Fig.2, while the participants were re-colored by sites.

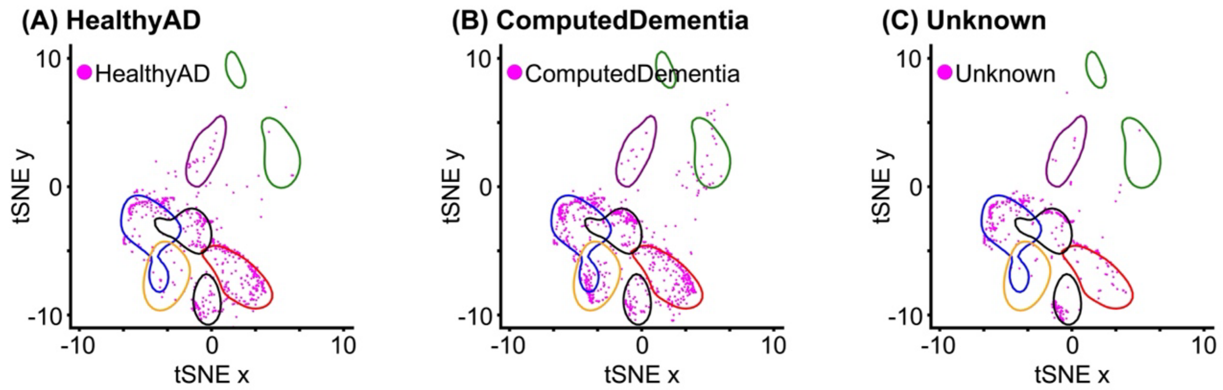


**Figure S3. Projection of held-out test participants onto the fitted 2-dimensional tSNE diagnostic probability map.** Test participants were excluded from model training and development. (A) HealthyAD group, participants clinically diagnosed with Alzheimer's disease but with preserved cognition (MMSE ≥ 26); (B) ComputedDementia group, participants without an available clinical diagnosis but with impaired cognition (MMSE < 19 or CDR ≥ 1); (C) Unknown group, participants with no available clinical diagnosis or cognitive scores (MMSE/CDR).

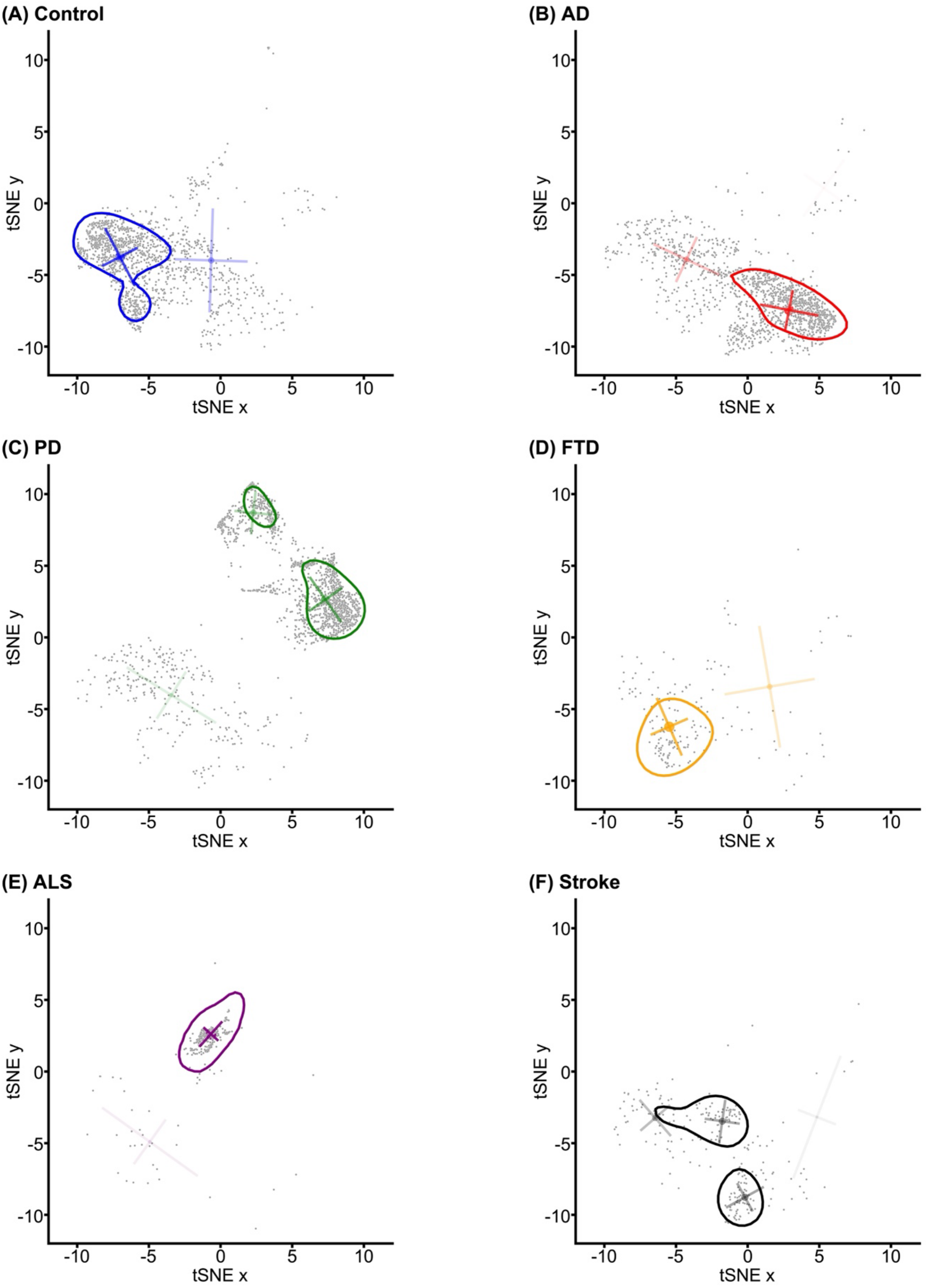


**Figure S4. K-Means clustering of participants by diagnosis on the tSNE diagnostic probability map.** The contour was drawn based on Gaussian kernel density estimation with a threshold of 0.01. (A) Control participants have two clusters. (B) AD patients have three clusters. (C) PD patients have four clusters. (D) FTD patients have two clusters. (E) ALS participants have two clusters. (F) Stroke/TIA participants have five clusters.

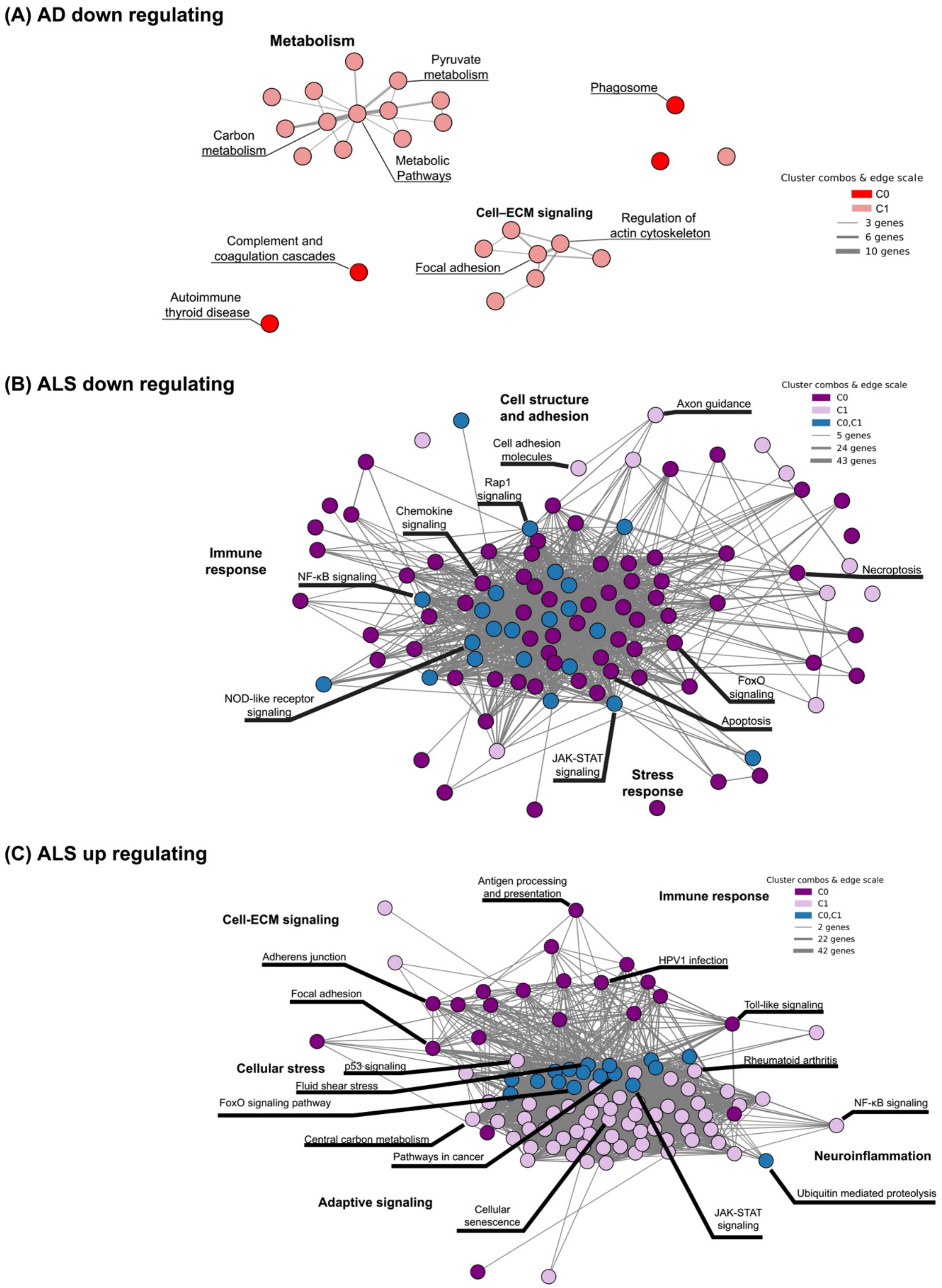


**Figure S5. KEGG terms (pFDR < 0.05) enriched among up- or down-regulated proteins in AD and ALS clusters.** Node shape denotes KEGG community; color indicates cluster. (A) KEGG terms enriched among down-regulated proteins across three AD clusters; (B) KEGG terms enriched among down-regulated proteins across two ALS clusters; (C) KEGG terms enriched among up-regulated proteins across two ALS clusters.

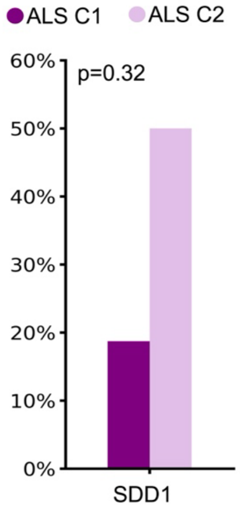


**Figure S6. The distribution of SOD1 mutations varied across the ALS clusters.** Among patients with available SOD1 information, 3/16 (18.8%) in ALS cluster C1 and 1/2 (50.0%) in ALS cluster C2 harbored SOD1 mutations. P value was unadjusted.

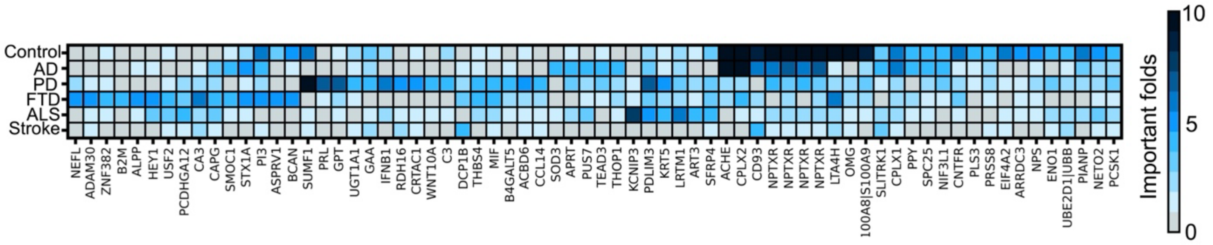


**Figure S7. Predictive importance of proteins by fold counts with MCI-SCI patients in the model development.** Proteins were visualized if they showed significant feature importance in more than 4 folds for at least one of the diagnostic tasks.

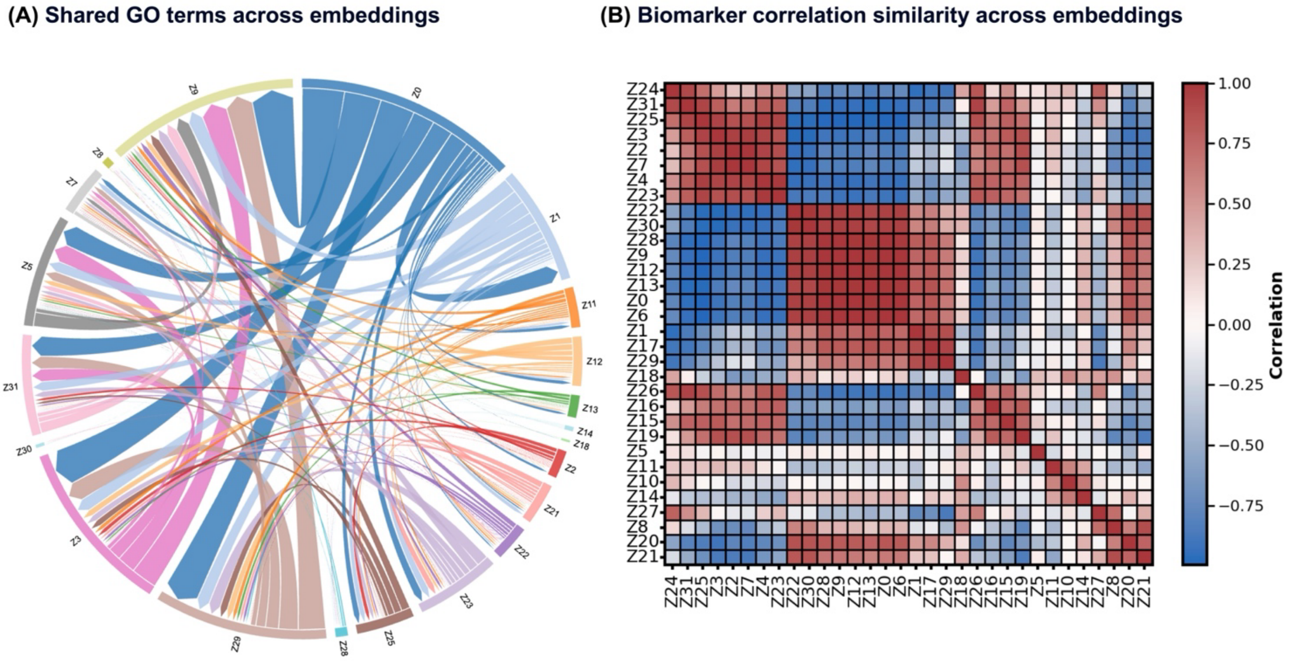


**Figure S8. Global view of embeddings.** (A) Network of embeddings connected by shared Gene Ontology (GO) terms. Nodes represent individual embeddings; edges indicate overlap in GO-term annotations, and edge width is proportional to the number of shared GO terms between the connected embeddings (larger width = greater overlap). (B) Similarity of embeddings based on biomarker-correlation profiles. For each embedding, we computed its correlation with the 21 biomarkers shown in Fig. 4C, yielding a 21-element correlation vector per embedding; pairwise embedding similarities were then calculated as the correlation between these correlation vectors.

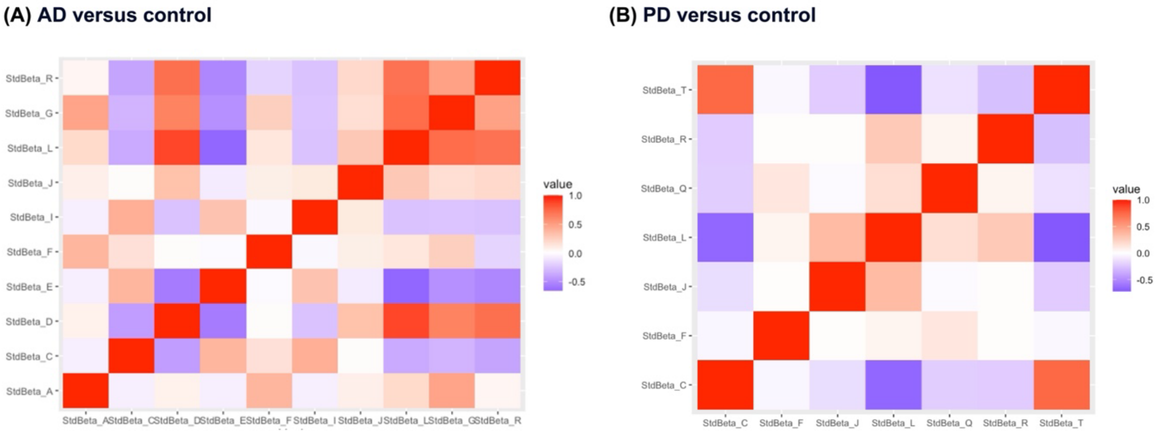


**Figure S9. Correlation of individual-protein effect sizes between sites.** At each site, a univariate logistic regression was run for every protein to examine the effect of disease status (disease vs control), with age, sex, and mean standardized protein level included as covariates; reported effect sizes correspond to the regression coefficients (β). For each pair of sites, we computed the similarity between sites by correlating their per-protein β vectors. The heatmap (or scatterplot) shows pairwise Pearson correlation values. (A) AD versus control. (B) PD versus control.

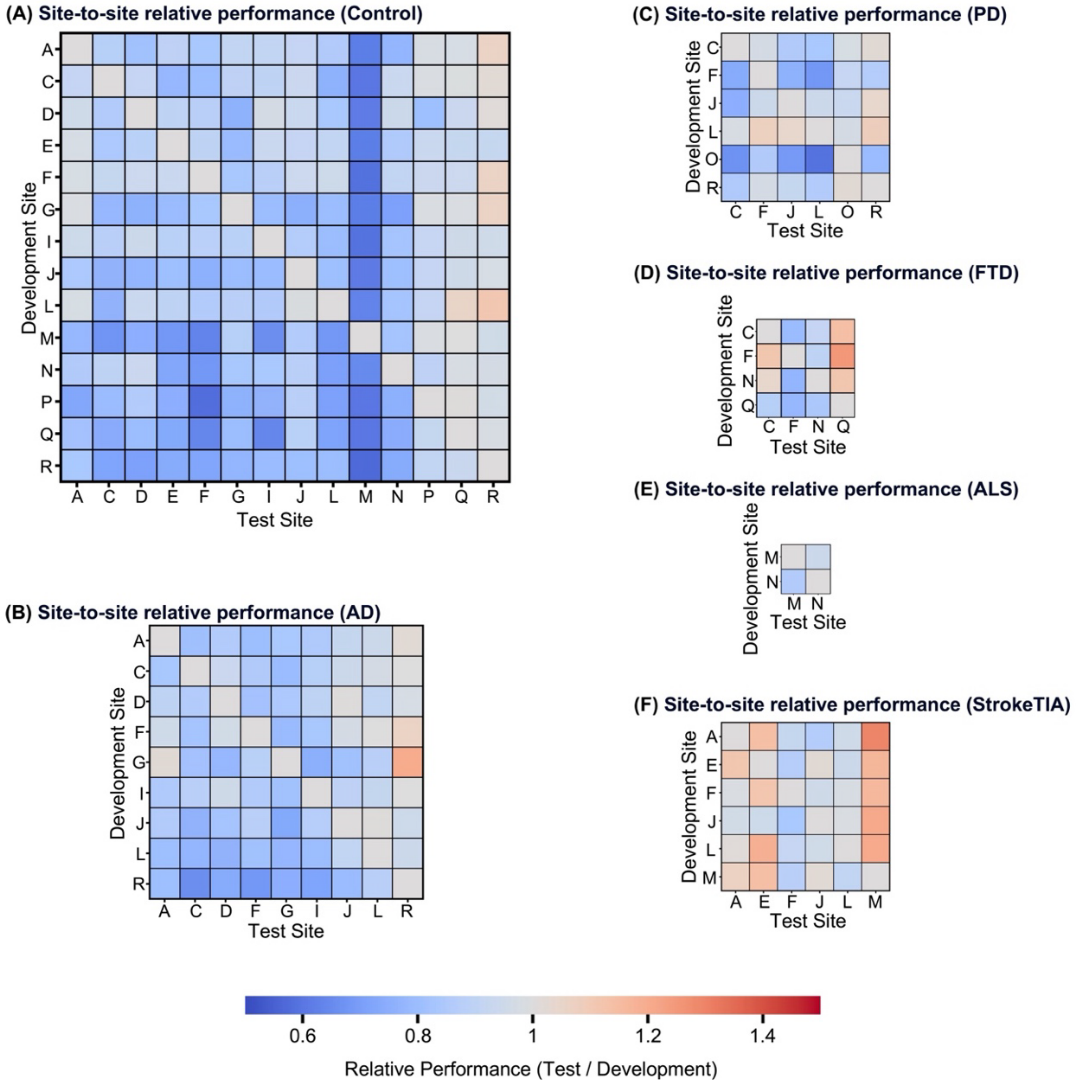


**Figure S10. Site-to-site generalization performance using TabPFN.** Heatmaps show the change in model performance when a TabPFN model trained on a development site is evaluated on a separate test site. Blue tones indicate poorer generalization; red tones indicate improved performance on the test site. (A) to (F) performances for Control, AD, PD, FTD, ALS, stroke/TIA respectively.

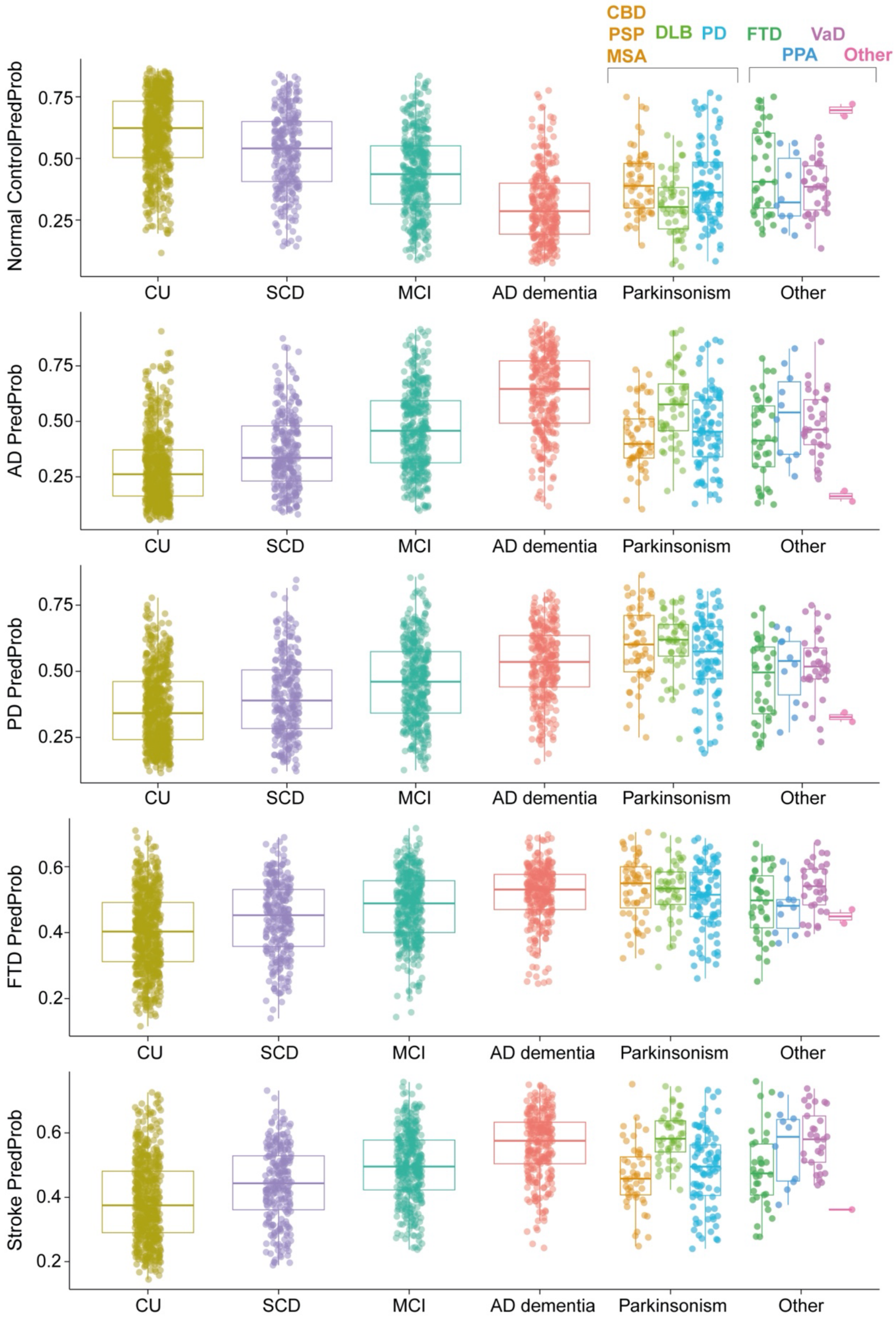


**Figure S11. Probabilities by each diagnostic group on the BioFINDER-2 cohort.**

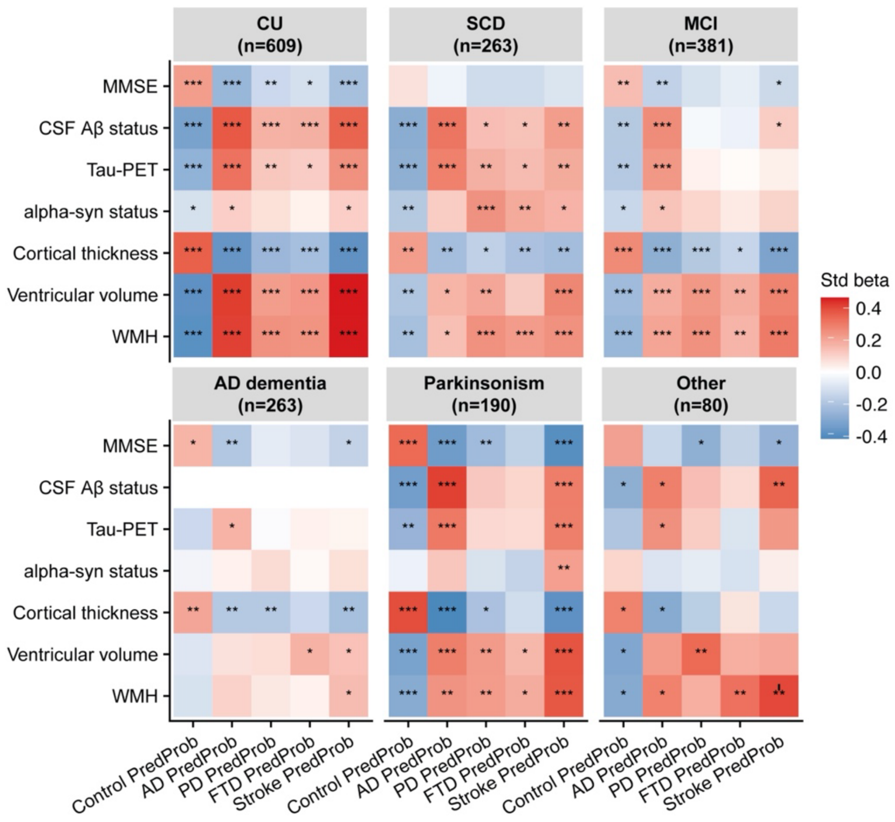


**Figure S12. Correlation between predicted probabilities and biomarkers.** Significant p values after FDR corrections across all comparisons are annotated with *.

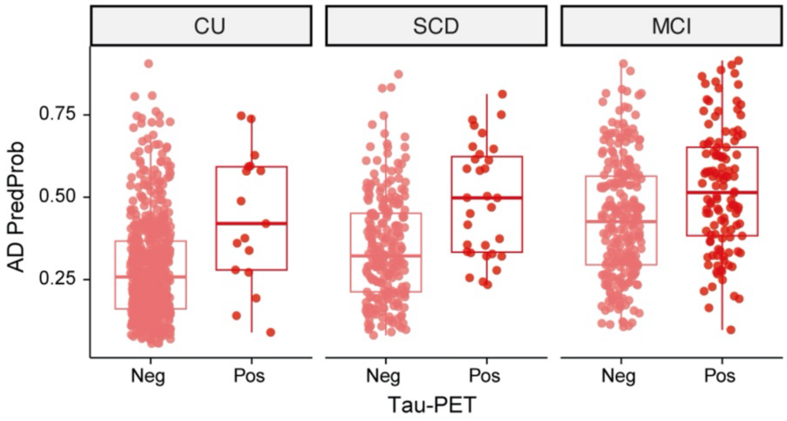


**Figure S13. AD probabilities by TauPET positivity across diagnostic groups on the BioFINDER-2 cohort.**

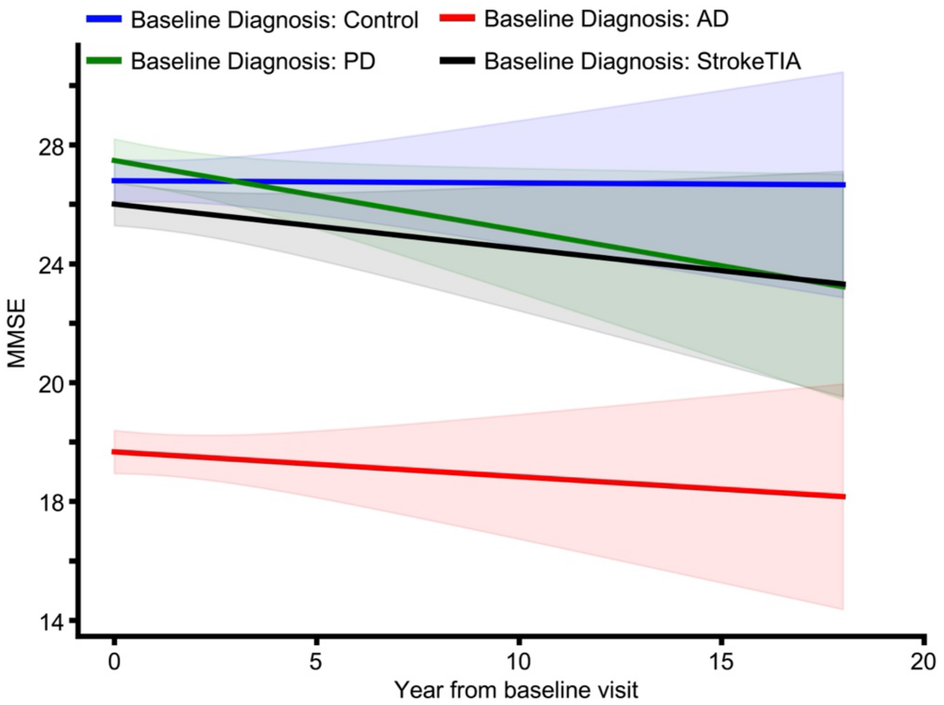


**Figure S14. MMSE longitudinal trajectories by baseline clinical diagnosis on the GNPC.** Longitudinal trajectories were assessed using the linear mixed-effects model: MMSE ~ Age + Sex + Site + BaselineDx × Year + (Year | SubjectID). Fixed effects: Age, Sex, Site, and the interaction between baseline diagnosis and time (Year); random intercepts and slopes for Year were included for each subject. The BaselineDx × Year term tests whether the annual change in MMSE differs by baseline diagnosis. Shaded bands = 95% CI.

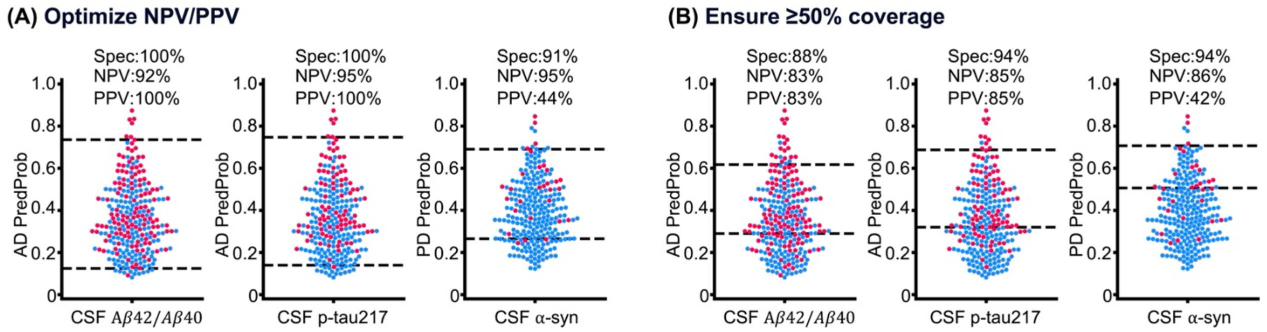


**Figure S15. Two cutoff strategies for predicting biomarker positivity.** Cutoffs were fit in non-SCD participants from BioFINDER-2 and then applied to individuals with SCD to estimate out-of-sample predictive performance. (A) Cutoffs optimized to maximize positive predictive value (PPV) and negative predictive value (NPV). (B) Cutoffs constrained to achieve ≥50% coverage.

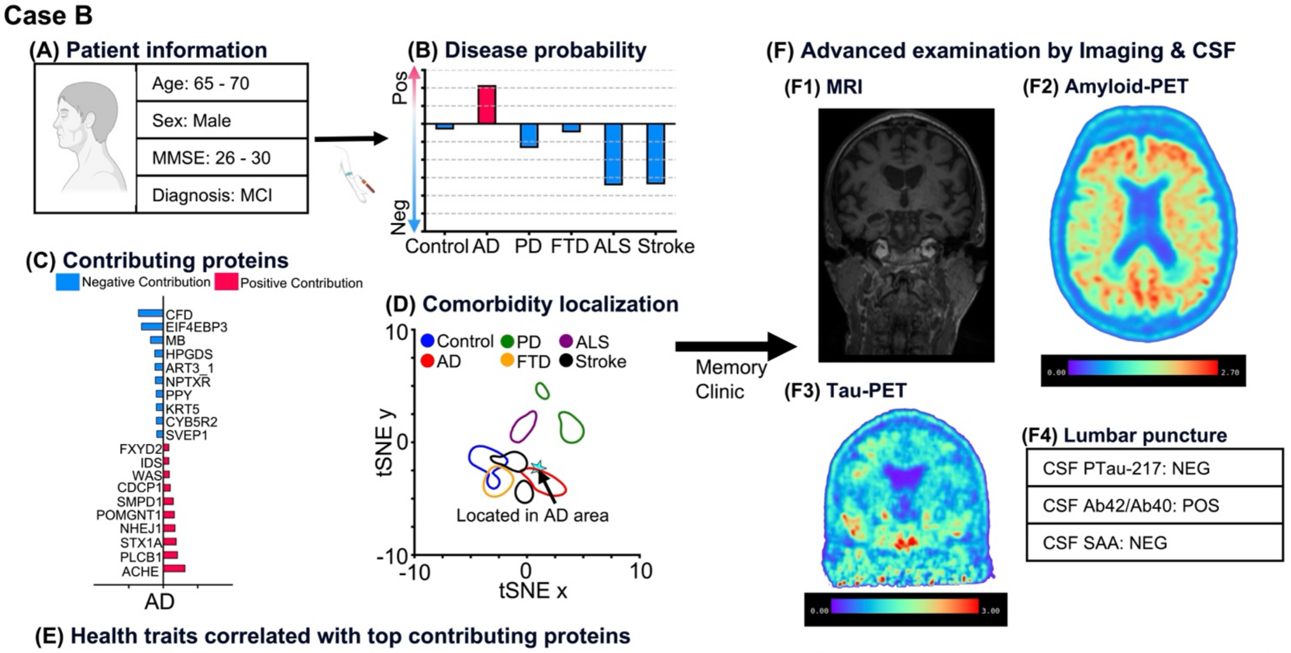


**(E) Health traits correlated with top contributing proteins**

| Protein | Interpretation | Positively Associated Traits | Negatively Associated Traits |
|---|---|---|---|
| ACHE | Higher AD Probability | Aspartate aminotransferase, Alanine aminotransferase | Platelet count, Platelet crit |
| CFD | Lower AD Probability | BMI, Creatinine | Docosahexaenoic Acid to Total Fatty Acids percentage, Docosahexaenoic Acid |

**Figure S16. Individual neurodegeneration risk report (Case B).** (A) Demographics and cognition score of one older participant, who was diagnosed with MCI. (B) ProtAIDe-Dx predicted this participant with higher probabilities of AD. The probabilities were normalized to be centered at zero. (C) Contributing proteins for making the decision were computed based on SHAP values. The top 10 positive and top 10 negative proteins were visualized. (D) The location of this participant on the diagnostic probability map indicates he was close to typical AD patients. (E) Health traits correlated with top contributing proteins were listed, informing clinicians to pay attention to these traits. (F) Advanced imaging and CSF biomarker examination at the memory clinic to confirm AD pathology.

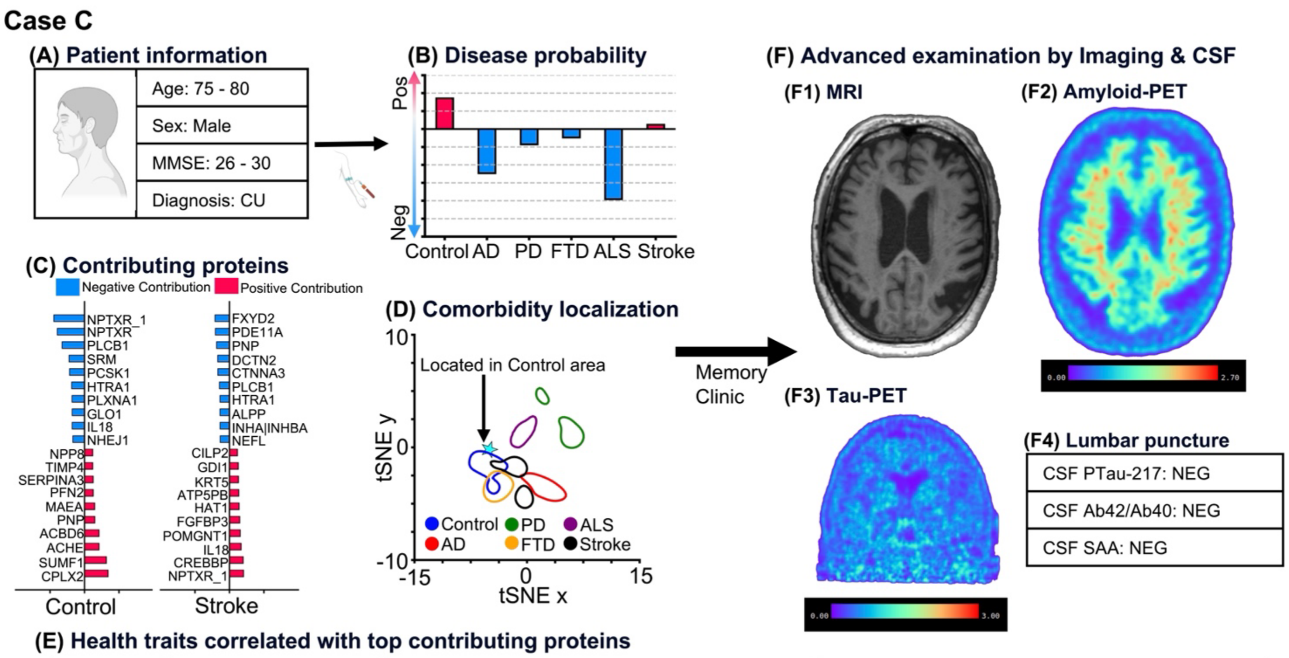


**(E) Health traits correlated with top contributing proteins**

| Protein | Interpretation | Positively Associated Traits | Negatively Associated Traits |
|---|---|---|---|
| CPLX2 | Higher Control probability | Creatinine, Cystatin C | BMI, Valine |
| NPTXR_1 | Lower Control probability<br>Higher Stroke probability | Cystatin C, Creatinine | BMI, Leucine |
| FXYD2 | Lower Stroke probability | Lactatem, Platelet count | Mean platelet (thrombocyte) volume, Glutamine |

**Figure S17. Individual neurodegeneration risk report (Case C).** (A) Demographics and cognition score of one older participant, who was diagnosed with CU. (B) ProtAIDe-Dx predicted this participant with higher probabilities of Control and stroke/TIA. The probabilities were normalized to be centered at zero. (C) Contributing proteins for making the decision were computed based on SHAP values. The top 10 positive and top 10 negative proteins were visualized. (D) The location of this participant on the diagnostic probability map indicates he was close to typical Stroke/TIA patients. (E) Health traits correlated with top contributing proteins were listed, informing clinicians to pay attention to these traits. (F) Advanced imaging and CSF biomarker examination at the memory clinic to confirm stroke/TIA pathology.

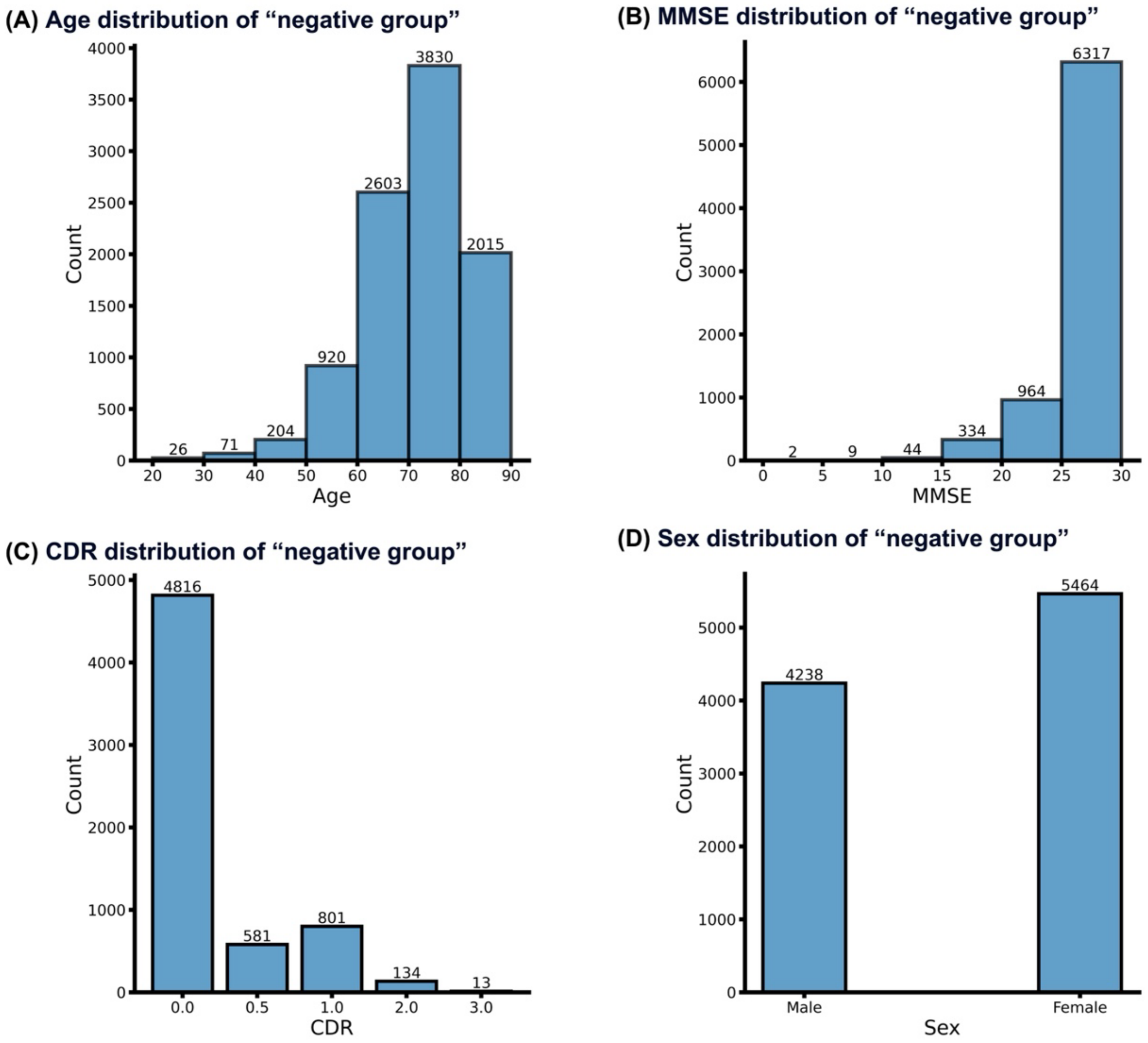


**Figure S18. Distribution of "negative group" (N=9,708)**. (A) Age distribution of "negative group". (B) MMSE distribution of "negative group". (C) CDR distribution of "negative group". (D) Sex distribution of "negative group".

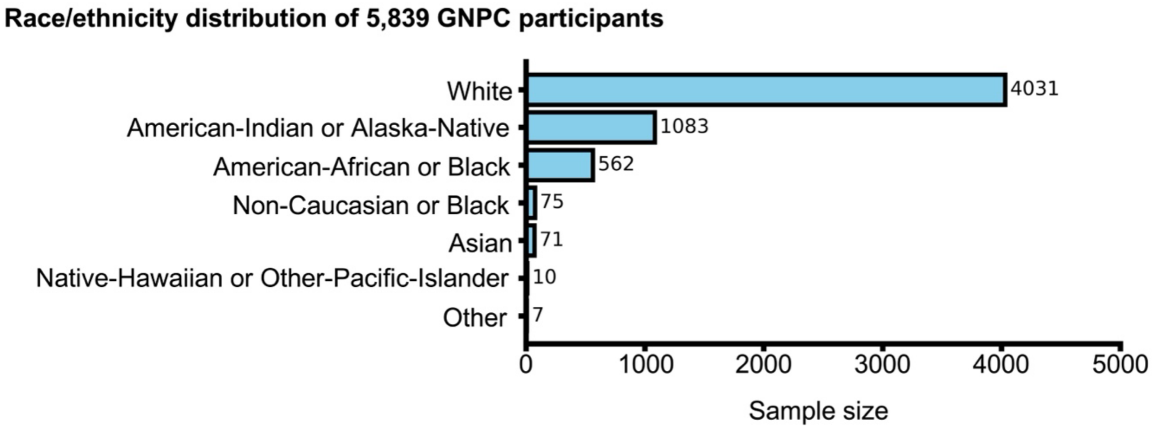


**Figure S19. Distribution of race and ethnicity among 5,839 GNPC participants.** As race and ethnicity were not primary variables during the GNPC collection stage, this information was available for only 5,839 participants.

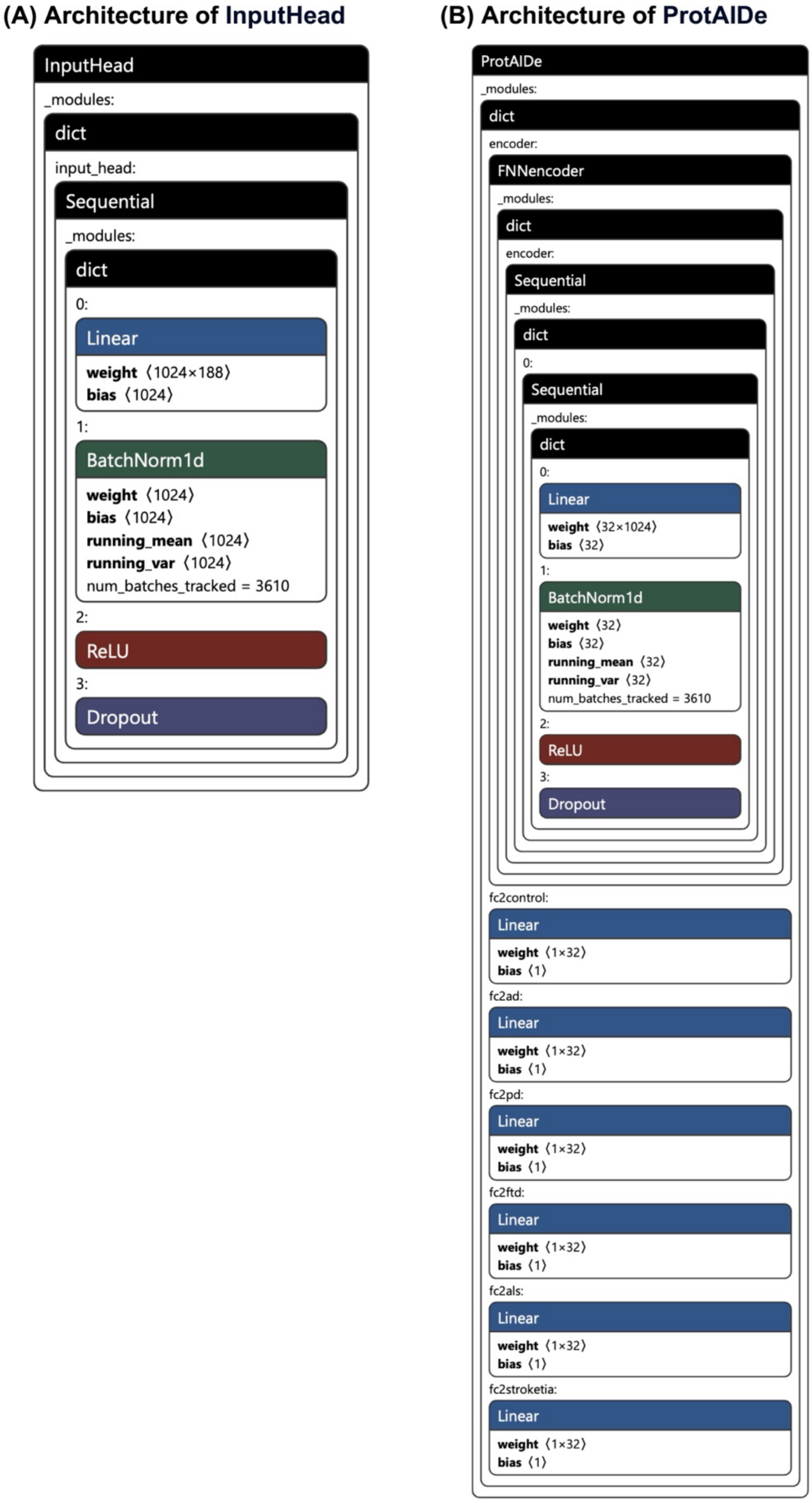


**Figure S20. Architecture of ProtAIDe-Dx (model evaluated on the BioFINDER-2 cohort).** The model comprises two MLP-based modules: (A) InputHead, which ingests proteins and produces initial input embeddings; and (B) ProtAIDe, which refines input embeddings and outputs probabilistic predictions for six conditions. Network components: Linear (affine transformation), BatchNorm1D (batch normalization over the feature dimension), ReLU (rectified linear unit), and Dropout (randomly zeroing activations with probability p). All ProtAIDe-Dx models share this architecture but differ in hyperparameters (e.g., number of layers, node of layer, dropout rate).

### Supplementary Tables

| Site | Sample Size | Age (Mean ± Std) | Sex (%Female) | BMI (Mean ± Std) | MMSE (Mean ± Std) |
|---|---|---|---|---|---|
| A | 983 | 72.1±6.7 | 56.4% | 27.9±5.6 | 26.6±2.9 |
| B | 443 | 57.0±4.9 | 60.2% | 26.8±4.1 | 29.1±1.0 |
| C | 2000 | 68.8±11.7 | 48.8% | 25.6±4.0 | 26.8±3.8 |
| D | 489 | 71.3±9.6 | 60.1% | - | 25.5±5.5 |
| E | 537 | 72.6±7.2 | 57.2% | 27.4±4.8 | 27.5±3.6 |
| F | 3355 | 74.4±10.0 | 56.7% | 27.5±5.3 | 26.3±4.6 |
| G | 1330 | 67.3±14.0 | 52.3% | - | 28.5±1.7 |
| I | 545 | 66.6±7.8 | 58.7% | 28.4±5.7 | - |
| J | 421 | 71.3±8.4 | 51.5% | 26.0±4.7 | - |
| K | 406 | 71.8±5.5 | 50.4% | 26.2±4.3 | 19.8±1.1 |
| L | 1101 | 78.8±8.8 | 58.9% | 26.4±5.2 | 26.2±5.1 |
| M | 355 | 57.6±12.0 | 43.9% | 27.3±5.0 | - |
| N | 789 | 56.1±14.7 | - | 26.9±5.8 | - |
| P | 519 | 71.7±8.7 | 44.5% | - | 28.3±1.4 |
| Q | 1370 | 72.2±8.8 | 57.9% | 26.9±4.1 | 24.5±4.5 |
| R | 435 | 82.7±5.7 | 71.0% | - | 22.7±8.2 |
| S | 200 | 60.0±9.8 | 50.0% | - | - |
| T | 233 | 66.5±8.3 | 65.2% | 26.4±4.5 | 28.2±1.7 |
| U | 1676 | 67.7±9.1 | 34.8% | - | - |

**Table S1. Demographics and cognition distribution of the selected 17,187 GNPC v1.3MS participants by site.** The item with "-" means this variable is missing.

| Site | Sample Size | %Control | %AD | %PD | %FTD | %ALS | %Stroke | %TIA |
|---|---|---|---|---|---|---|---|---|
| A | 983 | 38.6% | 26.2% | 0.1% | 0 | 0 | 3.2% | 5.1% |
| B | 443 | 80.8% | 0 | 0 | 0 | 0 | 0 | 0 |
| C | 2000 | 36.2% | 13.2% | 6.4% | 0 | 0 | 0 | 0 |
| D | 489 | 28.4% | 55.8% | 0 | 0 | 0 | 0 | 0 |
| E | 537 | 57.5% | 10.8% | 0.4% | 0.7% | 0 | 0.4% | 2.2% |
| F | 3355 | 46.5% | 29.2% | 3.5% | 0.9% | 0 | 2.6% | 0 |
| G | 1330 | 69.7% | 10.5% | 0 | 0 | 0 | 1.1% | 0 |
| I | 545 | 42.9% | 35.2% | 0.2% | 2.8% | 0 | 0 | 0.7% |
| J | 421 | 42.3% | 22.1% | 24.0% | 0 | 0 | 0.7% | 2.6% |
| K | 406 | 0 | 0 | 0 | 0 | 0 | 0 | 0 |
| L | 1101 | 47.1% | 11.8% | 6.2% | 0 | 0 | 10.7% | 11.3% |
| M | 355 | 30.1% | 0 | 0 | 0.3% | 69.0% | 1.7% | 3.1% |
| N | 789 | 48.7% | 0.4% | 0.1% | 12.5% | 1.1% | 0 | 0 |
| P | 519 | 86.9% | 0 | 0 | 0 | 0 | 1.5% | 0 |
| Q | 1370 | 25.0% | 0.3% | 3.9% | 0.7% | 0 | 0 | 0 |
| R | 435 | 26.4% | 31.0% | 4.8% | 0 | 0 | 23.4% | 0 |
| S | 200 | 0 | 0 | 0 | 0 | 95.0% | 0 | 0 |
| T | 233 | 0 | 0 | 100% | 0 | 0 | 0 | 0 |
| U | 1676 | 0 | 0 | 100% | 0 | 0 | 0 | 0 |

**Table S2. Positive ratio of clinical diagnosis of the selected 17,187 GNPC v1.3MS.** The item with "-" means this variable is missing, and the item with 0 means all participants are negative. The disease diagnostic categories were not exclusive, and some participants missed diagnostic information for six conditions, therefore, the sum of positive ratios was not equal to 1.

| Metric | Model | **Control** mean±std median | **AD** mean±std median | **PD** mean±std median | **FTD** mean±std median | **ALS** mean±std median | **Stroke** mean±std median |
|---|---|---|---|---|---|---|---|
| BCA | RandomForest | 0.81±0.01 0.80 | 0.65±0.02 0.65 | 0.89±0.01 0.89 | 0.50±0 0.50 | 0.81±0.03 0.82 | 0.54±0.02 0.54 |
| | XGBoost | 0.83±0.01 0.83 | 0.78±0.02 0.78 | 0.92±0.01 0.92 | 0.55±0.03 0.55 | 0.95±0.02 0.94 | 0.60±0.03 0.61 |
| | TabPFN | 0.84±0.01 0.84 | 0.82±0.02 0.82 | 0.92±0.01 0.92 | 0.64±0.06 0.63 | 0.95±0.04 0.96 | 0.70±0.04 0.70 |
| | ProtAIDe-Dx | 0.83±0.01 0.83 | 0.82±0.02 0.81 | 0.92±0.01 0.92 | 0.72±0.04 0.74 | 0.95±0.03 0.95 | 0.69±0.03 0.70 |
| | Ensemble | 0.85±0.01 0.84 | 0.82±0.01 0.82 | 0.92±0.01 0.93 | 0.73±0.03 0.74 | 0.95±0.03 0.96 | 0.70±0.03 0.71 |
| AUC | RandomForest | 0.81±0.01 0.80 | 0.65±0.02 0.65 | 0.89±0.01 0.89 | 0.50±0 0.50 | 0.81±0.03 0.82 | 0.54±0.02 0.54 |
| | XGBoost | 0.92±0.01 0.91 | 0.91±0.01 0.91 | 0.97±0.01 0.97 | 0.84±0.02 0.84 | 0.99±0.01 1.00 | 0.78±0.04 0.79 |
| | TabPFN | 0.92±0.01 0.92 | 0.92±0.01 0.92 | 0.97±0.01 0.97 | 0.86±0.04 0.86 | 0.96±0.00 1.00 | 0.82±0.03 0.82 |
| | ProtAIDe-Dx | 0.92±0.01 0.92 | 0.91±0.01 0.90 | 0.97±0.01 0.97 | 0.86±0.04 0.87 | 0.99±0.01 1.00 | 0.79±0.03 0.78 |
| | Ensemble | 0.93±0.01 0.92 | 0.91±0.01 0.91 | 0.97±0.01 0.97 | 0.86±0.04 0.87 | 0.99±0.01 1.00 | 0.79±0.03 0.78 |

**Table S3. BCA and AUC scores for 10-fold cross-validation**. Mean, standard deviation, and median values were computed from 10 test folds.

| Metric | Model | **Control** mean±std median | **AD** mean±std median | **PD** mean±std median | **FTD** mean±std median | **ALS** mean±std median | **Stroke** mean±std median |
|---|---|---|---|---|---|---|---|
| BCA | RandomForest | 0.59±0.09 0.60 | 0.56±0.03 0.56 | 0.51±0.02 0.50 | 0.50±0 0.50 | 0.58±0.08 0.58 | 0.50±0.01 0.50 |
| | XGBoost | 0.64±0.07 0.64 | 0.67±0.05 0.67 | 0.58±0.04 0.57 | 0.50±0 0.50 | 0.56±0.04 0.56 | 0.51±0.01 0.50 |
| | TabPFN | 0.66±0.09 0.68 | 0.73±0.06 0.71 | 0.60±0.06 0.59 | 0.52±0.02 0.51 | 0.61±0.10 0.61 | 0.54±0.05 0.53 |
| | ProtAIDe-Dx | 0.67±0.09 0.68 | 0.73±0.06 0.74 | 0.62±0.07 0.60 | 0.69±0.04 0.68 | 0.56±0.06 0.56 | 0.59±0.06 0.59 |
| | Ensemble | 0.66±0.09 0.68 | 0.72±0.04 0.71 | 0.62±0.07 0.60 | 0.66±0.09 0.65 | 0.56±0.05 0.56 | 0.57±0.06 0.57 |
| | Retrain | 0.61±0.05 0.63 | 0.64±0.04 0.64 | 0.61±0.07 0.60 | 0.61±0.08 0.62 | 0.56±0.06 0.56 | 0.53±0.03 0.52 |
| | Finetune | 0.68±0.08 0.71 | 0.74±0.05 0.72 | 0.64±0.07 0.64 | 0.62±0.05 0.61 | 0.60±0.00 0.60 | 0.56±0.05 0.57 |
| AUC | RandomForest | 0.59±0.09 0.60 | 0.56±0.03 0.56 | 0.51±0.02 0.50 | 0.50±0 0.50 | 0.58±0.08 0.58 | 0.50±0.01 0.50 |
| | XGBoost | 0.72±0.08 0.74 | 0.80±0.07 0.83 | 0.71±0.10 0.73 | 0.63±0.06 0.64 | 0.74±0.02 0.74 | 0.57±0.07 0.57 |
| | TabPFN | 0.75±0.09 0.79 | 0.83±0.06 0.85 | 0.76±0.12 0.77 | 0.66±0.04 0.64 | 0.77±0.02 0.77 | 0.61±0.08 0.60 |
| | ProtAIDe-Dx | 0.74±0.08 0.78 | 0.81±0.06 0.82 | 0.71±0.09 0.69 | 0.72±0.04 0.72 | 0.66±0.03 0.66 | 0.61±0.02 0.62 |
| | Ensemble | 0.75±0.10 0.78 | 0.81±0.06 0.83 | 0.72±0.09 0.74 | 0.72±0.03 0.71 | 0.72±0.04 0.72 | 0.61±0.04 0.62 |
| | Retrain | 0.68±0.08 0.70 | 0.72±0.07 0.73 | 0.67±0.09 0.69 | 0.65±0.13 0.67 | 0.69±0.06 0.69 | 0.57±0.06 0.55 |
| | Finetune | 0.73±0.10 0.76 | 0.80±0.07 0.80 | 0.70±0.08 0.70 | 0.67±0.05 0.65 | 0.63±0.00 0.63 | 0.59±0.09 0.60 |

**Table S4. BCA and AUC scores for leave-one-site-out**. Mean, standard deviation, and median values were computed from 14 test sites.

| Target | Relative Performances (Test / Development) |
|---|---|
| Control | 84%±12% |
| AD | 87%±10% |
| PD | 89%±13% |
| FTD | 96%±16% |
| ALS | 90%±4% |
| Stroke | 100%±12% |

**Table S5. Mean and standard deviation of relative performances from site-to-site generalization experiment.** The 14 test sites were the same as in the leave-one-site-out setting.

| Variable | True Negative | False Positive | Adjusted P Values |
|---|---|---|---|
| Age | 60.4±15.7 | 78.6±6.7 | 4e-38 |
| Sex (% Female) | 60.1% | 56.7% | 0.59 |
| CSF abnormal Aβ42/Aβ40 ratio | 14.8% | 44.8% | 1e-8 |
| CSF p-tau 217 | 6.27±6.73 | 13.2±12.9 | 6e-5 |
| TauPET | 1.08±0.11 | 1.19±0.20 | 3e-5 |
| ADSignCT | 2.76±0.12 | 2.67±0.13 | 8e-7 |
| WholeBrainCT | 2.40±0.08 | 2.32±0.07 | 2e-10 |
| Ventricle Volume | 0.009±0.006 | 0.014±0.001 | 1e-8 |
| WMH | 0.003±0.003 | 0.006±0.004 | 3e-7 |

**Table S6. Distribution of demographics and AD biomarkers between the True Negative and False Positive groups on the BioFIDNER-2 cohort.** The true positive group (N=459) referred to Control participants predicted as Control, and the false positive group (N=67) referred to Control participants predicted as AD patients. For binary variables, p values were computed by the two-proportion z test; for continuous variables, p values were computed by the t-test. All p values were FDR corrected.

| Model | AD | PD | FTD | Stroke |
|---|---|---|---|---|
| M0 | 0.60±0.02 | 0.57±0.03 | 0.51±0.03 | 0.54±0.05 |
| M1 | 0.63±0.02 | 0.62±0.03 | 0.54±0.03 | 0.67±0.06 |
| M2 | 0.84±0.02 | 0.67±0.03 | 0.72±0.04 | 0.60±0.06 |
| M3 | 0.84±0.02 | 0.76±0.03 | 0.81±0.04 | 0.63±0.06 |

**Table S7. Models' BCA (mean±std) on differentiating diagnoses on the BioFINDER-2 cohort.** The mean and standard deviations were computed on 1000 bootstraps of testing participants.

| Model Pair | | FDR-corrected P values | | | |
|---|---|---|---|---|---|
| **Model A** | **Model B** | **AD** | **PD** | **FTD** | **Stroke** |
| M0 | M1 | 2e-118 | 4e-296 | 5e-64 | 0 |
| M0 | M2 | 0 | 0 | 8e-190 | 9e-30 |
| M0 | M3 | 0 | 0 | 0 | 9e-79 |
| M1 | M2 | 0 | 0 | 0 | 4e-212 |
| M1 | M3 | 0 | 2e-17 | 0 | 9e-22 |
| M2 | M3 | 0 | 0 | 0 | 5e-124 |

**Table S8. FDR-corrected p values by comparing the models' BCA on differentiating diagnoses on the BioFINDER-2 dataset.** The p values were computed on 1000 bootstraps with t-test. The BCA was defined as one-versus-rest.

| | Coef | Std.Err | Z | P value | Adjusted P value |
|---|---|---|---|---|---|
| Intercept | 19.73 | 0.37 | 53.33 | 4.72e-01 | 6.12e-01 |
| C(Sex) | -0.06 | 0.09 | -0.72 | 9.30e-01 | 9.30e-01 |
| C(Site)[D] | 0.02 | 0.22 | 0.09 | 1.31e-02 | 2.61e-02 |
| C(Site)[E] | 0.56 | 0.23 | 2.48 | 1.22e-02 | 2.61e-02 |
| C(Site)[F] | 0.40 | 0.16 | 2.51 | 6.39e-01 | 7.20e-01 |
| C(Site)[P] | -0.06 | 0.13 | -0.47 | 8.92e-04 | 2.38e-03 |
| C(Site)[R] | -1.05 | 0.31 | -3.32 | 8.92e-02 | 1.59e-01 |
| C(Site)[T] | -0.99 | 0.58 | -1.70 | 9.44e-141 | 7.55e-140 |
| C(BlDx)[Control] | 9.13 | 0.36 | 25.26 | 2.22e-49 | 8.89e-49 |
| C(BlDx)[PD] | 9.81 | 0.66 | 14.77 | 5.01e-50 | 2.67e-49 |
| C(BlDx)[StrokeTIA] | 8.34 | 0.56 | 14.87 | 4.36e-09 | 1.40e-08 |
| Age | -0.36 | 0.06 | -5.87 | 4.54e-01 | 6.12e-01 |
| Year | -0.08 | 0.11 | -0.75 | 4.97e-01 | 6.12e-01 |
| C(BlDx)[Control]:Year | 0.08 | 0.11 | 0.68 | 2.33e-01 | 3.73e-01 |
| C(BlDx)[PD]:Year | -0.15 | 0.13 | -1.19 | 6.75e-01 | 7.20e-01 |
| C(BlDx)[StrokeTIA]:Year | -0.07 | 0.16 | -0.42 | 4.72e-01 | 6.12e-01 |

**Table S9. Linear mixed effect model results for modeling cognitive decline by baseline diagnosis on GNPC.** MMSE ~ Age + Sex + Site + BaselineDx * Year + Year|SubjID. BlDx was short for BaselineDx. Fixed effects: Age, Sex, Site and the interaction between baseline diagnosis and time (Year); random intercepts and slopes for Year were included for each subject. The BaselineDx × Year term tests whether annual change in MMSE differs by baseline diagnosis.

| | Coef | Std.Err | Z | P value | Adjusted P value |
|---|---|---|---|---|---|
| Intercept | 19.22 | 0.40 | 47.74 | 0 | 0 |
| C(Sex) | -0.08 | 0.09 | -0.93 | 3.51e-01 | 4.76e-01 |
| C(Site)[D] | 0.13 | 0.22 | 0.60 | 5.48e-01 | 6.80e-01 |
| C(Site)[E] | 0.54 | 0.22 | 2.40 | 1.63e-02 | 2.81e-02 |
| C(Site)[F] | 0.41 | 0.16 | 2.57 | 1.01e-02 | 1.92e-02 |
| C(Site)[P] | -0.07 | 0.13 | -0.51 | 6.09e-01 | 6.80e-01 |
| C(Site)[R] | -0.57 | 0.54 | -1.06 | 2.88e-01 | 4.21e-01 |
| C(Site)[T] | -0.12 | 0.83 | -0.14 | 8.87e-01 | 8.87e-01 |
| C(BlDx)[Control] | 8.76 | 0.37 | 23.89 | 3.69e-126 | 3.51e-125 |
| C(BlDx)[PD] | 8.79 | 0.65 | 13.49 | 1.8e-41 | 8.82e-41 |
| C(BlDx)[StrokeTIA] | 7.83 | 0.54 | 14.43 | 3.13e-47 | 1.98e-46 |
| C(BlPred)[Control] | 0.92 | 0.33 | 2.81 | 4.96e-03 | 1.30e-02 |
| C(BlPred)[PD] | 0.38 | 0.73 | 0.52 | 6.05e-01 | 6.80e-01 |
| C(BlPred)[StrokeTIA] | 0.11 | 0.66 | 0.16 | 8.72e-01 | 8.87e-01 |
| Age | -0.34 | 0.06 | -5.51 | 3.65e-08 | 1.39e-07 |
| Year | -0.31 | 0.11 | -2.88 | 3.97e-03 | 1.26e-02 |
| C(BlPred)[Control]:Year | 0.29 | 0.11 | 2.74 | 6.18e-03 | 1.30e-02 |
| C(BlPred)[PD]:Year | 0.33 | 0.15 | 2.20 | 2.77e-02 | 4.39e-02 |
| C(BlPred)[StrokeTIA]:Year | 0.37 | 0.14 | 2.76 | 5.74e-03 | 1.30e-02 |

**Table S10. Linear mixed effect model results for modeling cognitive decline by baseline prediction on GNPC.** MMSE ~ Age + Sex + Site + BaselineDx + BaselinePrediction * Year + Year|SubjectID. BlDx was short for BaselineDx, and BlPred was short for BaselinePrediction. Fixed effects: Age, Sex, Site, BaselineDx, and the interaction between baseline prediction and time (Year); random intercepts and slopes for Year were included for each subject. The BaselinePrediction × Year term tests whether annual change in MMSE differs by baseline prediction.

| | Coef | Std.Err | Z | P value | Adjusted P value |
|---|---|---|---|---|---|
| Intercept | 28.43 | 1.17 | 24.21 | 1.59e-129 | 9.57e-129 |
| C(Sex) | -0.55 | 0.24 | -2.28 | 2.28e-02 | 2.73e-02 |
| C(BlPred)[Control] | 0.80 | 0.26 | 3.12 | 1.80e-03 | 2.70e-03 |
| Age | -0.02 | 0.02 | -1.23 | 2.17e-01 | 2.17e-01 |
| Year | -1.62 | 0.15 | -10.65 | 1.80e-26 | 5.41e-26 |
| C(BlPred)[Control]:Year | 0.73 | 0.22 | 3.37 | 7.48e-04 | 1.50e-03 |

**Table S11. Linear mixed effect model results for modeling cognitive decline by baseline prediction on BioFINDEER-2 MCI patients.** MMSE ~ Age + Sex + BaselinePrediction * Year + Year| SubjectID. BlPred was short for BaselinePrediction. Fixed effects: Age, Sex, Site, and the interaction between baseline prediction and time (Year); random intercepts and slopes for Year were included for each subject. The BaselinePrediction × Year term tests whether annual change in MMSE differs by baseline prediction.

| Diagnosis Group | Sample size | Age (Mean ± Std) | Sex (n,%Female) | MMSE (Mean ± Std) |
|---|---|---|---|---|
| CU | 609 | 63.6±15.8 | 350 (57.4%) | 29.0±1.1 |
| SCD | 263 | 68.0±9.0 | 133 (50.6%) | 28.7±1.5 |
| MCI | 381 | 71.7±7.9 | 162 (42.5%) | 27.1±1.9 |
| AD dementia | 263 | 73.8±7.4 | 147 (55.9%) | 21.0±4.2 |
| Parkinsonism | 190 | 71.0±8.8 | 52 (27.4%) | 25.8±4.5 |
| Other diseases | 80 | 72.0±8.8 | 36 (45.0%) | 24.0±4.1 |

**Table S12. Demographics and cognition distribution of 1,786 BioFINDER-2 participants.**

| Site | Control (Pos-Neg) | AD (Pos-Neg) | PD (Pos-Neg) | FTD (Pos-Neg) | ALS (Pos-Neg) | Stroke/TIA (Pos-Neg) |
|---|---|---|---|---|---|---|
| A | 379-250 | 214-412 | X | X | X | 57-572 |
| C | 723-399 | 224-898 | 127-995 | 48-1074 | X | X |
| D | 139-126 | 125-76 | X | X | X | X |
| E | 309-76 | X | X | X | X | 14-313 |
| F | 1527-780 | 690-1617 | 99-2208 | 25-2282 | X | 74-2233 |
| G | 927-40 | 26-941 | X | X | X | X |
| I | 234-206 | 192-133 | X | X | X | X |
| J | 178-201 | 93-285 | 101-278 | X | X | 14-347 |
| L | 519-308 | 113-128 | 65-283 | X | X | 192-510 |
| M | 107-248 | X | X | X | 245-110 | 11-344 |
| N | 384-112 | X | X | 99-397 | 9-487 | X |
| P | 451-8 | X | X | X | X | X |
| Q | 343-66 | X | 54-355 | 10-399 | X | X |
| R | 115-191 | 126-180 | 20-285 | X | X | X |

**Table S13. Sample size by each target for selected testing sites in GNPC.** The ‘X’ means the target of this contributor did not pass the selection criteria.

| Hyperparameter | Range |
|---|---|
| Node of input layer | 16, 32, 64, 128, 256, 512, 1024 |
| Number of hidden layers | 1, 2, 3 |
| Node of each hidden layer | 8, 16, 32, 64, 128, 256, 512 |
| Learning rate | 1e-4 – 1e-1 |
| Dropout | 0 - 0.5 |
| Optimizer | Adam, SGD, RMSProp |
| $\lambda$ | 0 - 5 |
| $\alpha$ | 0 - 0.2 |

**Table S14. Hyperparameter search range for ProtAIDe-Dx model.**

| Hyperparameter | Optimal value |
|---|---|
| Node of input layer | 1024 |
| Number of hidden layers | 1 |
| Node of each hidden layer | 32 |
| Learning rate | 1e-4 |
| Dropout | 0.4 |
| Optimizer | Adam |
| $\lambda$ | 3 |
| $\alpha_{Control}$ | 0.1 |
| $\alpha_{AD}$ | 0.1 |
| $\alpha_{PD}$ | 0.1 |
| $\alpha_{FTD}$ | 0.1 |
| $\alpha_{ALS}$ | 0 |
| $\alpha_{stroke/TIA}$ | 0.1 |

**Table S15. Optimal hyperparameters of the ProtAIDe-Dx model evaluated on the BioFINDER-2 cohort.**
